# Simulating drug effects on blood glucose laboratory test time series with a conditional WGAN

**DOI:** 10.1101/2020.07.19.20157321

**Authors:** Alexandre Yahi, Nicholas P. Tatonetti

## Abstract

The unexpected effects of medications has led to more than 14 million drug adverse events reported to the Food and Drug Administration (FDA) over the past 10 years in the United States alone, with a little over 1.3 million of them linked to death, and represents a medical and financial burden on our healthcare. Laboratory tests have the potential to capture inter-individual variability in drug responses, but a significant portion of the patient population has unique treatment pathways that impedes forecasting and optimal decision making.

Generative Adversarial Networks (GANs) are flexible implicit generative models that have demonstrated their ability to capture complex correlations in field like computer vision and natural language. Their latent representation capacity is an opportunity for drug effect simulation on laboratory test trajectories. In this paper, we developed and evaluated conditional GANs on glucose laboratory tests in patients exposed to drug combinations and showed a proof of concept for these models in the simulation of unseen drug combinations. By using conditional Wasserstein GANs (WGANs) to simulate drug effects in laboratory tests, we hope to pave the way for novel clinical decision support (CDM) systems and enable the development of better predictive models for rare cohorts of patients.

## 1 Introduction

Drug effects can be unpredictable.

Each novel therapeutics submitted for approval to the Food and Drug Administration (FDA) needs to be safe and effective for its intended use. However, FDA approval does not guarantee safety and effectiveness for all patients. In fact, the response rates of patients to most major drugs fall in the 50 to 75% range.[57] This is due to the variability in treatment response among patients, known as inter-patient variability, caused by factors such as the environment, genetics, polypharmacy or comorbidities [58, 17, 68]. The consequences for drug safety are more concerning: out of 222 novel therapeutics approved by the FDA between 2001 and 2010, 71 (32%) were flagged with post-market safety events, including 61 incremental boxed warnings for 43 of these therapeutics [18]. Between 2008 and 2017, the FDA approved 321 novel drugs. Over the same period of time, the FDA Adverse Event Reporting System (FAERS) recorded more than 10 million AE reports, among which 5.8 million were serious adverse drug reactions (SADRs), and 1.1 million were AEs related to death. AEs burden our health system causing 2 million hospital stays each year and lengthening visits by 1.7 to 4.6 days[2]. The economic, social, and health burden of these events make pharmacovigilance an essential and pressing public health concern.

The solution is to pick the right treatment for the right patient using all the resources available. While clinical trials try to ascertain that a drug is safe and effective for its intended use before its marketing, pharmacovigilance centers monitor AE reports and aim at ensuring that a drug’s safety information is up to date. However, neither of these processes are error proof. On one hand, clinical trials have focused on designing drugs for the average patient[60] even at a time when there are increasing calls for precision medicine to enable the ”right drug at the right dose to the right patient”[14]. On the other hand, spontaneous reporting systems are known to suffer from biases such as under-reporting which is especially troublesome for rare events and drug-drug interactions (DDIs)[39]. Therefore it is only by using post-marketing observational data that we can uncover off-label uses, treatment patterns, patient specific variability in responses, and rare ADRs [28, 51, 52, 53, 54, 55].

Unfortunately, a large number of patients have unique treatment trajectories that makes their outcomes hard to predict. In a large-scale EHR study, Hirpcsak et al. [29] showed that many patients have unique treatment pathways. They analyzed 11 EHR data sources that had adopted the Observational Health Data Sciences and Informatics (OHDSI) common data model (CDM), in four different countries and including 250 million patient records. By enumerating 3-year treatment pathways, they found that 10% of patients with type 2 diabetes, 11% of patients with depression, and 24% of patients with hypertension could not compare their treatment pathway with anyone else in this quarter billion population of diverse individuals.

Therefore, there is a need to go beyond patient matching and classic supervised machine learning models that suffer from unbalanced training classes or the sample size of population with rare events. We need methods to interpolate sets of conditional information unseen – or very rare – in the dataset of interest, to support decision making and biomedical predictive models.

With the development of novel deep learning methods such as generative adversarial networks (GANs), there is an opportunity to learn how to augment existing clinical datasets with realistic synthetic data and increase predictive performances. Moreover, GANs have the potential to simulate effects of individual covariates such as drug exposures by leveraging the properties of implicit generative models. Instead of modeling every covariate and confounding variable, conditional GANs can learn how to match auxiliary clinical information to conditionally learned distributions from which they can be stochastically sampled. Although the amount of information these models can retain naturally cannot exceed the information available in the input data, conditional models appear to disentangle information from other sample classes, and other combinations of auxiliary information to infer conditional distributions unseen at training time. In this paper, our contributions are two-fold:

1. we developed and evaluated deep implicit generative models to learn distributions of laboratory test time series using two versions of the WGAN: the WGAN with gradient penalty (WGAN-GP) [26], and the WGAN with Lipschitz penalty (WGAN-LP) [44];
2. we studied the use of conditional generative adversarial networks (GANs) to model laboratory test time series and demonstrate how these models can be used for the simulation of drug effects.

We provided two applications as proof of concepts of these proposed models. The first is to illustrate the latent representation power of these conditional GAN by showing that we can infer laboratory test time series associated with drug exposure combinations unseen at training. The second is its direction consequence: the data augmentation of rare events to improve their predictability. We conducted the targeted augmentation of the 10 rarest drug combinations occurring during glucose lab test trajectories and we were able to improve forecasting on seven of them by adding synthetic samples to real samples during training of predictive models. These are evidence that these methods should be explored further.

## 2 Methods

### 2.1 Data selection

All the data come from Columbia University Irving Medical Center/New York Presbyterian Hospital (CUIMC/NYPH) transformed for the Observational Medical Outcomes Partnership (OMOP) common data model (CDM) v5, including inpatient and outpatient records. CUIMC/NYPH is an academic medical center with over 1000 inpatient beds serving both adult and pediatric populations. The laboratories receive on average over 10,000 samples a day. Over 550 different assays are performed on-site in several laboratories, including Core (Hematology and Chemistry), Microbiology, Molecular Diagnosis, Immunogenetics, Cytogenetics, and several Specialty Laboratories and Satellite Laboratories. Over 15 million assays are performed annually in-house. Due to the complexity of the cases treated at our hospital, over 200,000 assays and panels are sent to over 60 different outside reference laboratories every year.[33]

As of 2019, we worked with a structured database of 6.38 million patients that counts:

- 78.95 million drug orders, representing 40.76 million single ingredient exposures, for 1.41 million patients,
- 140.30 million diagnosis codes for 5.40 million patients,
- 64.38 million procedure codes for 3.58 million patients,
- 810.68 million measurements (i.e., laboratory tests and vitals) for 2.29 million patients.

We considered inpatient visits as a unit of analysis, with about 38.49 million visit occurrences recorded in our research database. We mapped each visit to the measurements performed throughout its duration. Each laboratory test time series (LTTS) was therefore a time series of value from a given laboratory test or vital, for a given patient, during a unique visit. Limiting these time series to a visit versus considering time series for the whole patient medical history enabled us to have more reasonable time intervals and account for clinical events that are more relevant to the time series at hand. The general statistics of unique time series length available during these visits are displayed in table 1. That table has 3 main take aways: (1) vitals (i.e., respiratory rate, heart rate, blood pressure…) are the most abundant measurements available but do not represent measurements with the most time series. (2) the most abundant time series are for laboratory tests that belong to the routine blood panels; (3) extreme values show that we are dealing with data that can present errors when entered in the EHR or exported for research purpose, and quality control is required.

**Table 1:**
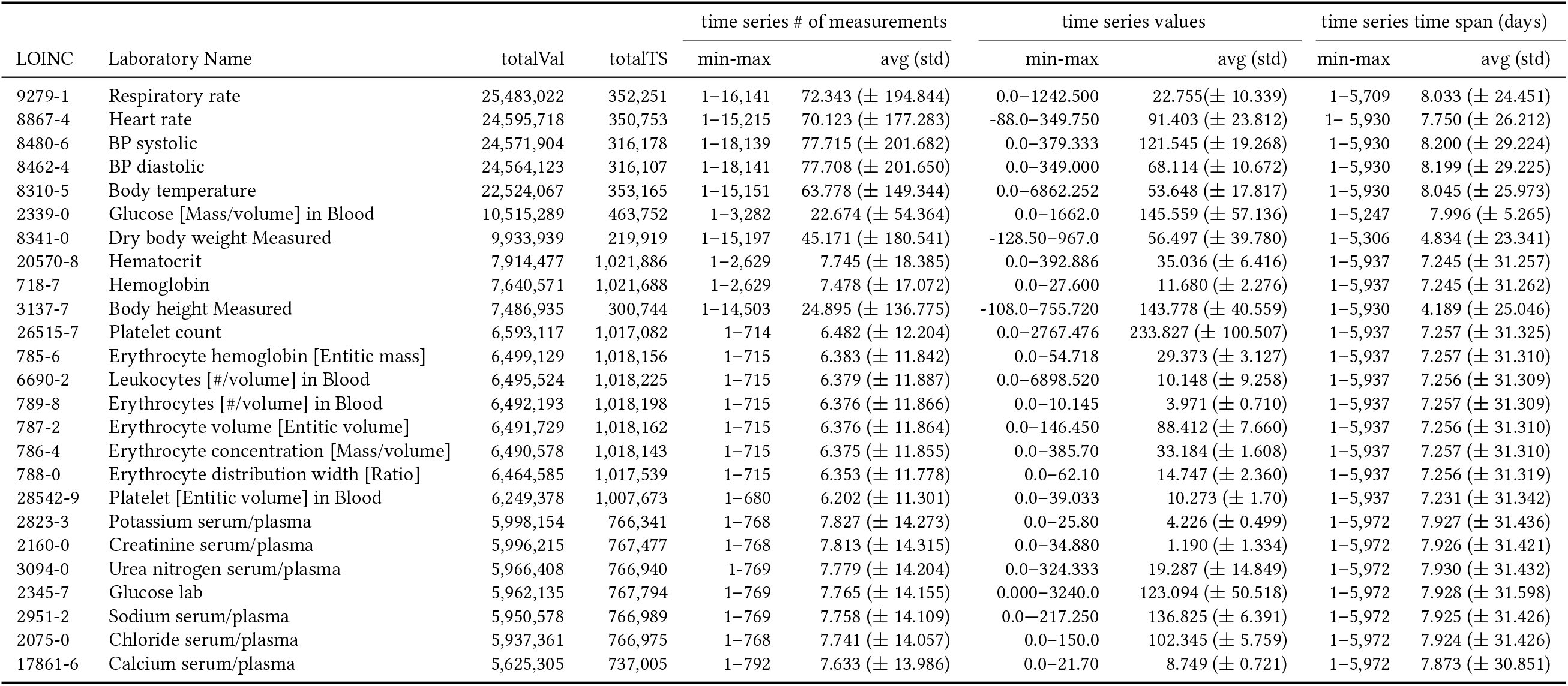
Summary statistics of the top-25 most recorded measurements at CUIMC/NYP; names have been edited for length; min-max ranges display the presence of outliers and errors in the source database.

We will now focus on the data specific to each modality: regularly sampled laboratory tests for the regular WGAN as a baseline, and irregularly sampled laboratory tests with drug exposures for the conditional WGAN.

#### 2.1.1 Regularly sampled time series

We restricted the sample set of laboratory test time series (LTTS) to the ones that have a regular sampling for the experiments with a non-conditional GAN. When multiple measurements were available the same day, we averaged them. We explored the amount of data available in function of the wanted length of time series (i.e., 5, 10 or 15 values) and the regular or irregular nature of sampling (Figure 1). We selected a time windows of 10 days which provided a good balance of number of features available for modeling and amount of samples. We observed that the longer the time series, the smaller the sample size, in particular for the most abundance measurements. When a given visit has multiple time series candidates available, we took the earliest for consistency.

**Figure 1:**
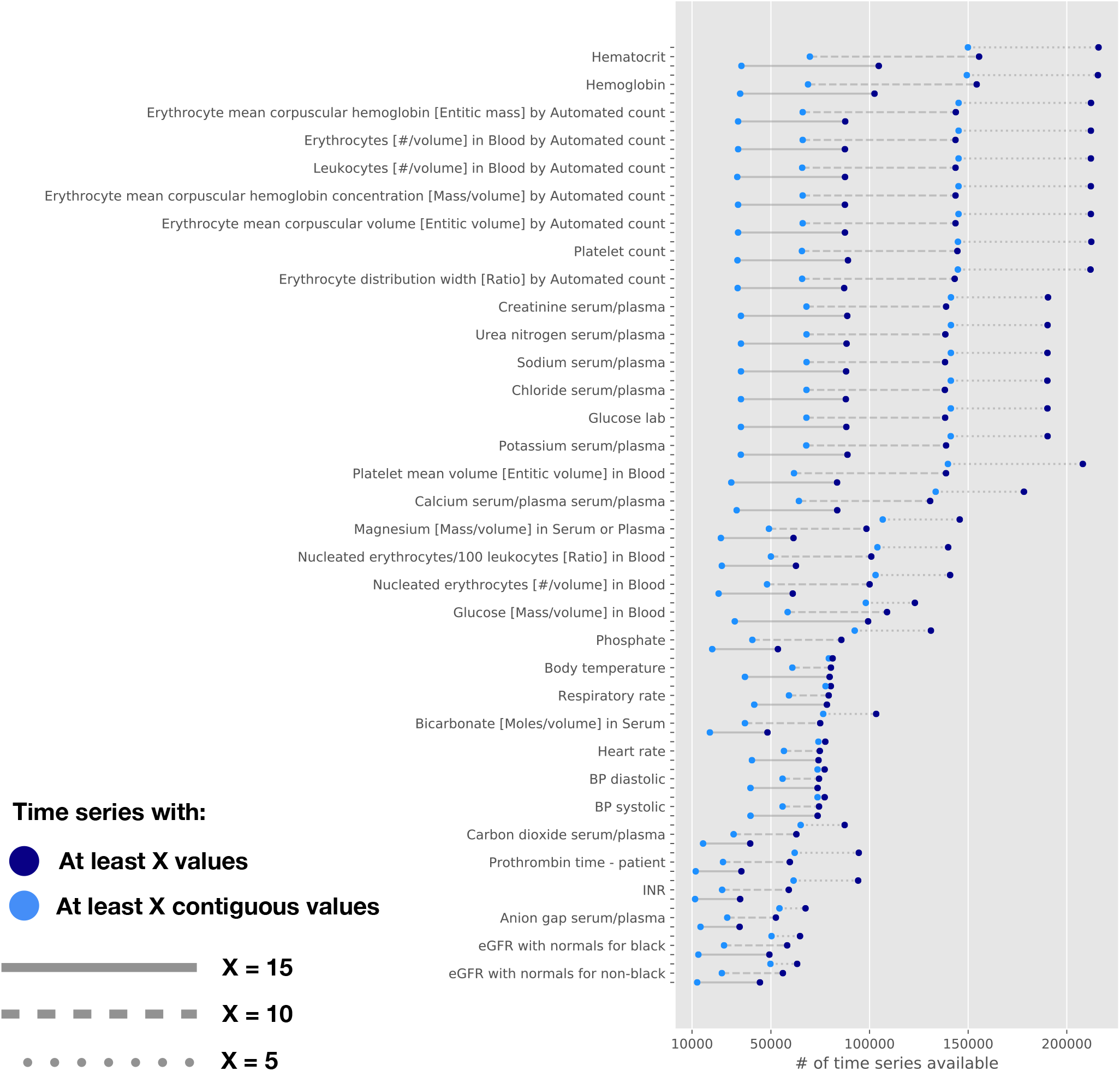
Laboratory test time series above 10,000 samples, for 3 different lengths, and 2 types of sampling.

The regularly sampled laboratory test time series restricted to a 10-day length are characterized in table 2. We computed the average dispersion (i.e., the ratio of the standard deviation by the mean of each time series) for all laboratory tests to get a sense of the relative variance of each measurement type. We selected glucose lab (LOINC 2345-7) as the main measurement to be modeled throughout this paper, for its high dispersion, satisfactory number of samples available, and link to various physiological processes.

**Table 2:** Summary statistics for top-20 laboratory tests when filtering for contiguous time series of length days=10. The laboratory test selected for modeling is highlighted in bold.

| LOINC | Laboratory Name | # time series | Dispersion (Std) |
| --- | --- | --- | --- |
| 2951-2 | Sodium serum/plasma | 85,626 | 0.032 (± 0.050) |
| 2075-0 | Chloride serum/plasma | 85,614 | 0.043 (± 0.048) |
| 3094-0 | Urea nitrogen serum/plasma | 85,600 | 0.273 (± 0.152) |
| 2160-0 | Creatinine serum/plasma | 85,586 | 0.183 (± 0.133) |
| **2345-7** | **Glucose lab** | **85,555** | **0.243 (± 0.141)** |
| 2823-3 | Potassium serum/plasma | 85,441 | 0.097 (± 0.053) |
| 20570-8 | Hematocrit | 84,056 | 0.092 (± 0.049) |
| 718-7 | Hemoglobin | 83,922 | 0.092 (± 0.048) |
| 789-8 | Erythrocytes [# /volume] in Blood | 82,909 | 0.090 (± 0.048) |
| 785-6 | Erythrocyte mean corpuscular hemoglobin [Entitic mass] | 82,892 | 0.019 (± 0.033) |
| 787-2 | Erythrocyte mean corpuscular volume [Entitic volume] | 82,880 | 0.020 (± 0.027) |
| 786-4 | Erythrocyte mean corpuscular hemoglobin concentration [Mass/volume] | 82,867 | 0.023 (± 0.029) |
| 6690-2 | Leukocytes [# /volume] in Blood | 82,729 | 0.266 (± 0.187) |
| 788-0 | Erythrocyte distribution width [Ratio] | 82,495 | 0.045 (± 0.093) |
| 26515-7 | Platelet count | 82,352 | 0.247 (± 0.167) |
| 17861-6 | Calcium serum/plasma serum/plasma | 80,667 | 0.053 (± 0.034) |
| 28542-9 | Platelet mean volume [Entitic volume] in Blood | 77,582 | 0.102 (± 0.289) |
| 2339-0 | Glucose [Mass/volume] in Blood | 62,774 | 0.192 (± 0.088) |
| 8310-5 | Body temperature | 62,766 | 0.053 (± 0.065) |
| 19048-8 | Nucleated erythrocytes/100 leukocytes [Ratio] in Blood | 62,231 | 0.931 (± 1.112) |

Before training models on these time series, we proceeded to a quality control step for all laboratory test to remove values outside of the 1-99 percentile range and remove extreme outliers or spurious values that could arise at the various steps of data collection and mapping. A time series with such values would be removed from the dataset.

We also computed the distribution of measurements for glucose lab in figure 2, a boxenplot to visualize the mean and percentiles in figure 3, along with a density heatmap of time series represented by their standard deviation and mean in figure 4.

**Figure 2:**
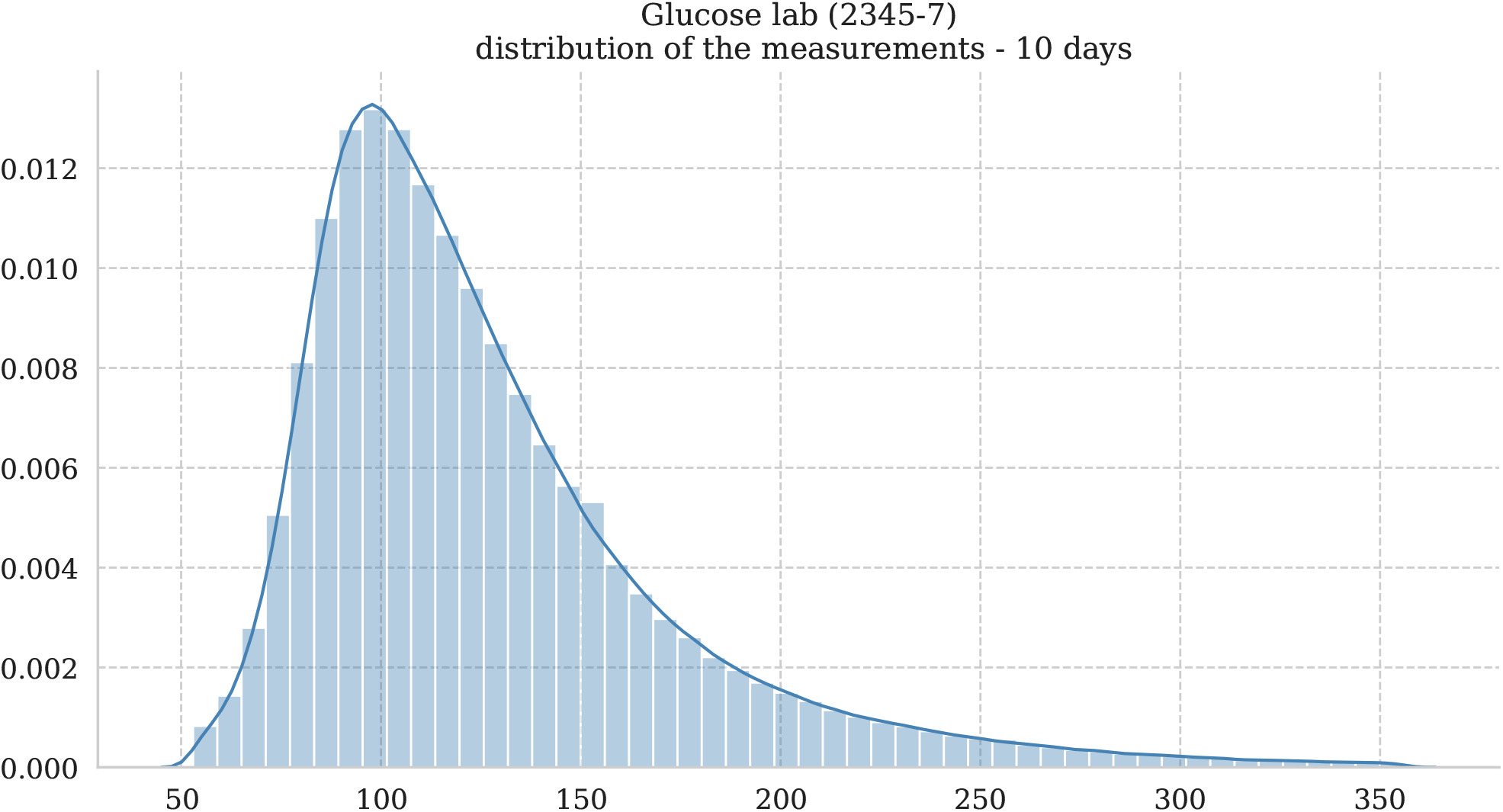
Distribution of glucose lab measurements post-quality control for regular time series.

**Figure 3:**
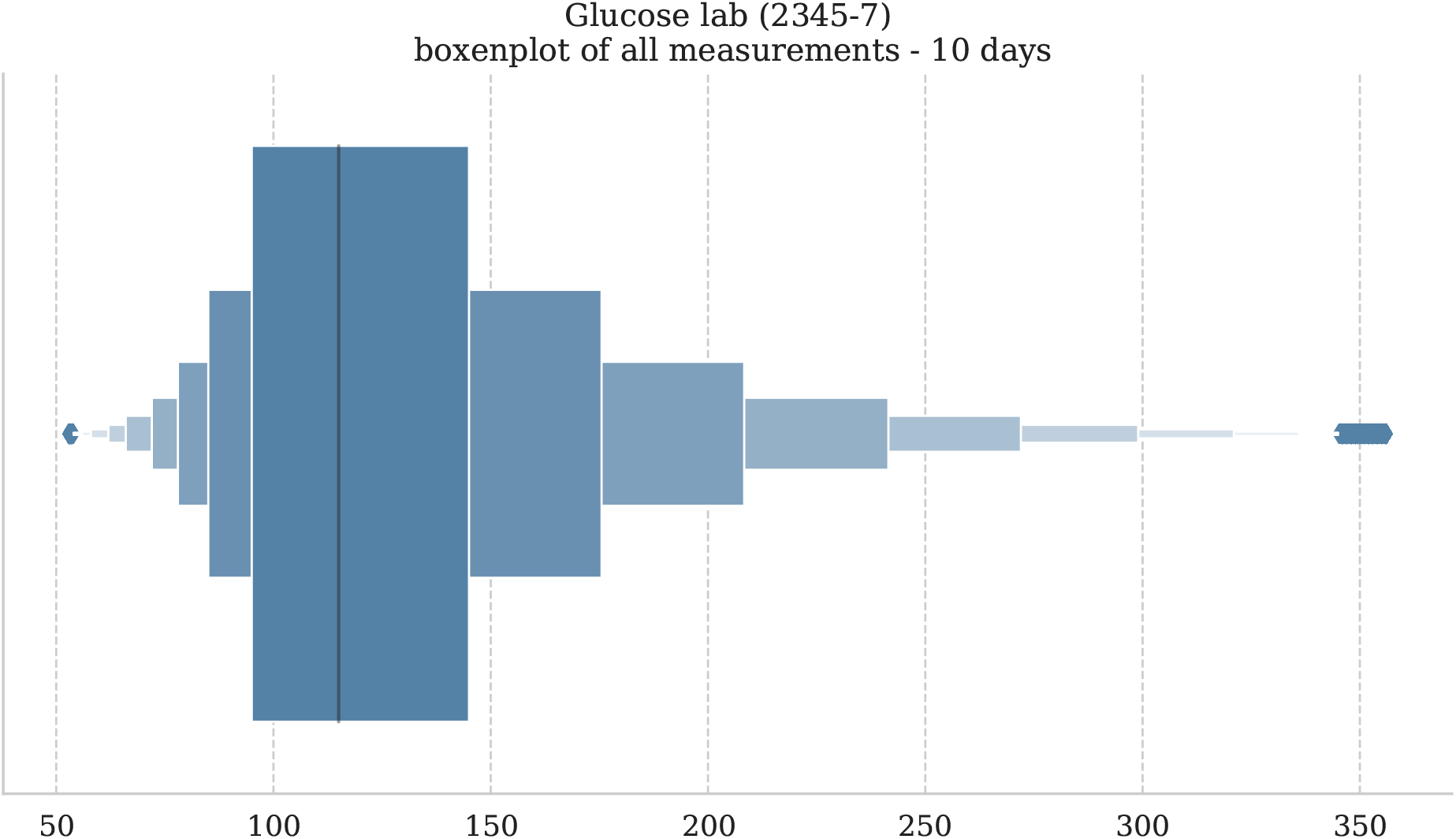
Boxenplot of glucose lab measurements post-quality control for regular time series to visualize percentiles and potential outliers.

**Figure 4:**
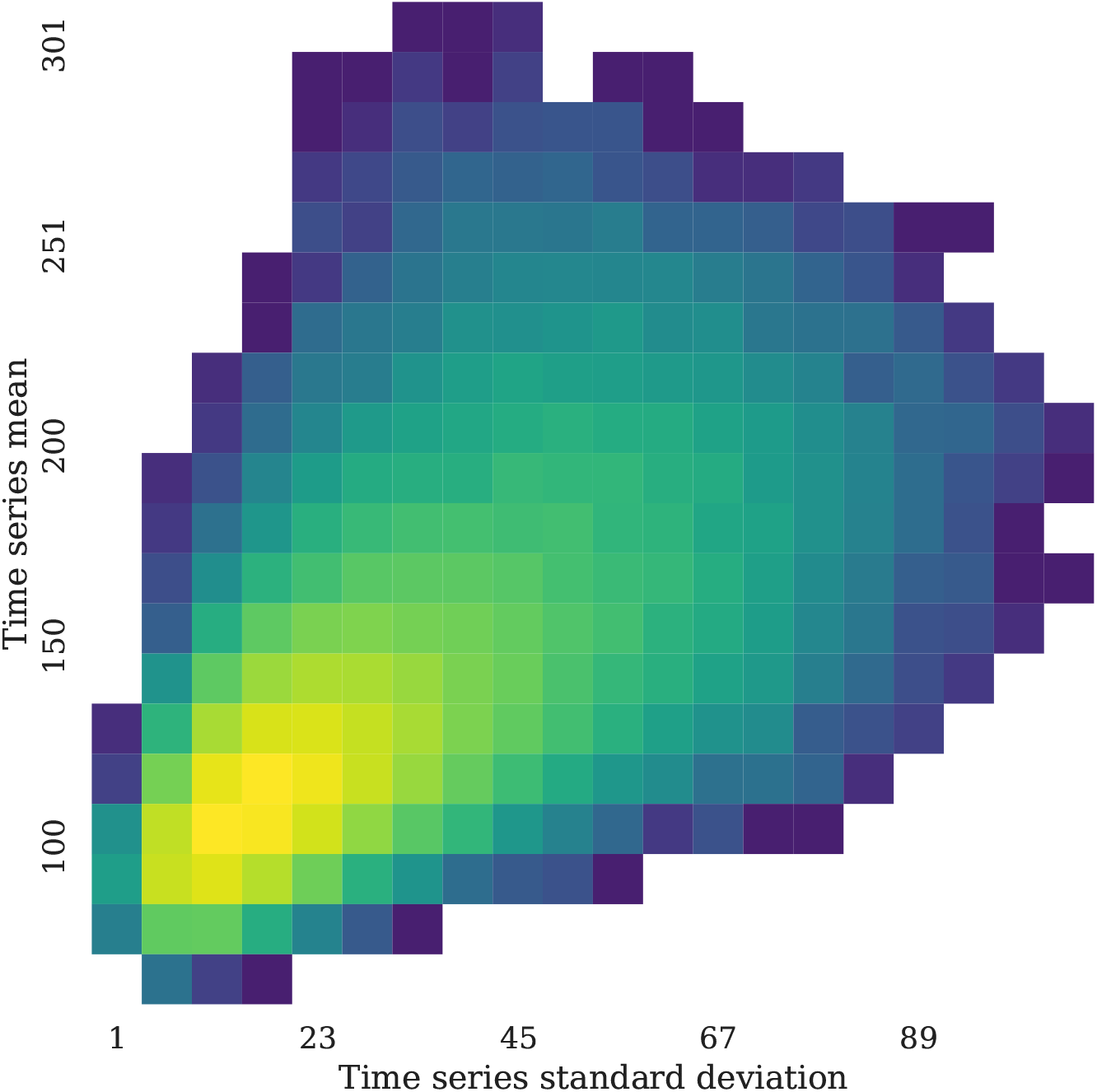
Density heatmap of regular glucose lab measurements time series represented by their standard deviations and mean.

#### 2.1.2 Irregularly sampled time series

For the conditional model, we lifted the constrain on regularity of the time series, to have a more realistic model. We used a forecasting task described in the *Supplementary Material* to evaluate if time information was relevant. Since time intervals were hurting the prediction of irregular time series, compared to models on regular time series with no time information, we focused on drug exposures as the sole auxiliary data type.

We selected time series of 10 values that can be spaced by more than one day, producing a set of irregularly sampled time series but with an constant number of measurements. In table 3 we can observe that relaxing the time interval constraints yields higher sample sizes.

**Table 3:** Summary statistics for top-20 irregularly sampled laboratory tests when filtering for total length=10. The top-5 laboratory tests with the highest dispersion index are highlighted.

| LOINC | Laboratory Name | # time series | Dispersion (Std) | Time interval (Std) |
| --- | --- | --- | --- | --- |
| 20570-8 | Hematocrit | 120,205 | 0.093 (± 0.051) | 1.271 (± 2.821) |
| 718-7 | Hemoglobin | 120,107 | 0.093 (± 0.051) | 1.271 (± 2.816) |
| 789-8 | Erythrocytes [# /volume] in Blood by Automated count | 119,278 | 0.091 (± 0.050) | 1.276 (± 2.839) |
| 785-6 | Erythrocyte mean corpuscular hemoglobin [Entitic mass] | 119,261 | 0.021 (± 0.037) | 1.276 (± 2.838) |
| 787-2 | Erythrocyte mean corpuscular volume [Entitic volume] | 119,254 | 0.022 (± 0.031) | 1.276 (± 2.839) |
| 786-4 | Erythrocyte mean corpuscular hemoglobin concentration [Mass/volume] | 119,251 | 0.023 (± 0.031) | 1.276 (± 2.838) |
| 6690-2 | Leukocytes [# /volume] in Blood | 119,243 | 0.264 (± 0.175) | 1.277 (± 2.839) |
| 788-0 | Erythrocyte distribution width [Ratio] | 119,007 | 0.047 (± 0.094) | 1.278 (± 2.842) |
| 26515-7 | Platelet count | 118,848 | 0.244 (± 0.165) | 1.278 (± 2.845) |
| 28542-9 | Platelet mean volume [Entitic volume] in Blood | 114,687 | 0.096 (± 0.266) | 1.309 (± 2.892) |
| 3094-0 | Urea nitrogen serum/plasma | 114,337 | 0.283 (± 0.156) | 1.190 (± 2.400) |
| 2160-0 | Creatinine serum/plasma | 114,329 | 0.184 (± 0.131) | 1.190 (± 2.406) |
| 2951-2 | Sodium serum/plasma | 114,324 | 0.031 (± 0.050) | 1.190 (± 2.430) |
| 2075-0 | Chloride serum/plasma | 114,303 | 0.042 (± 0.048) | 1.190 (± 2.435) |
| **2345-7** | **Glucose lab** | **114,248** | **0.248 (± 0.145)** | **1.188 (± 2.432)** |
| 2823-3 | Potassium serum/plasma | 114,144 | 0.100 (± 0.056) | 1.190 (± 2.437) |
| 17861-6 | Calcium serum/plasma serum/plasma | 107,956 | 0.054 (± 0.034) | 1.194 (± 2.360) |
| 19048-8 | Nucleated erythrocytes/100 leukocytes [Ratio] in Blood | 84,787 | 0.917 (± 1.114) | 1.233 (± 3.034) |
| 30392-5 | Nucleated erythrocytes [# /volume] in Blood | 83,879 | 0.711 (± 1.059) | 1.233 (± 2.717) |
| 19123-9 | Magnesium [Mass/volume] in Serum or Plasma | 81,994 | 0.107 (± 0.061) | 1.242 (± 1.569) |

After quality control, we summarized the among of time series left to train the models, along with their associated demographics in table 4. Figures 5,6 and 7 represent the distribution of measurements, boxenplot, and density heatmaps of the glucose lab irregular time series.

**Table 4:**
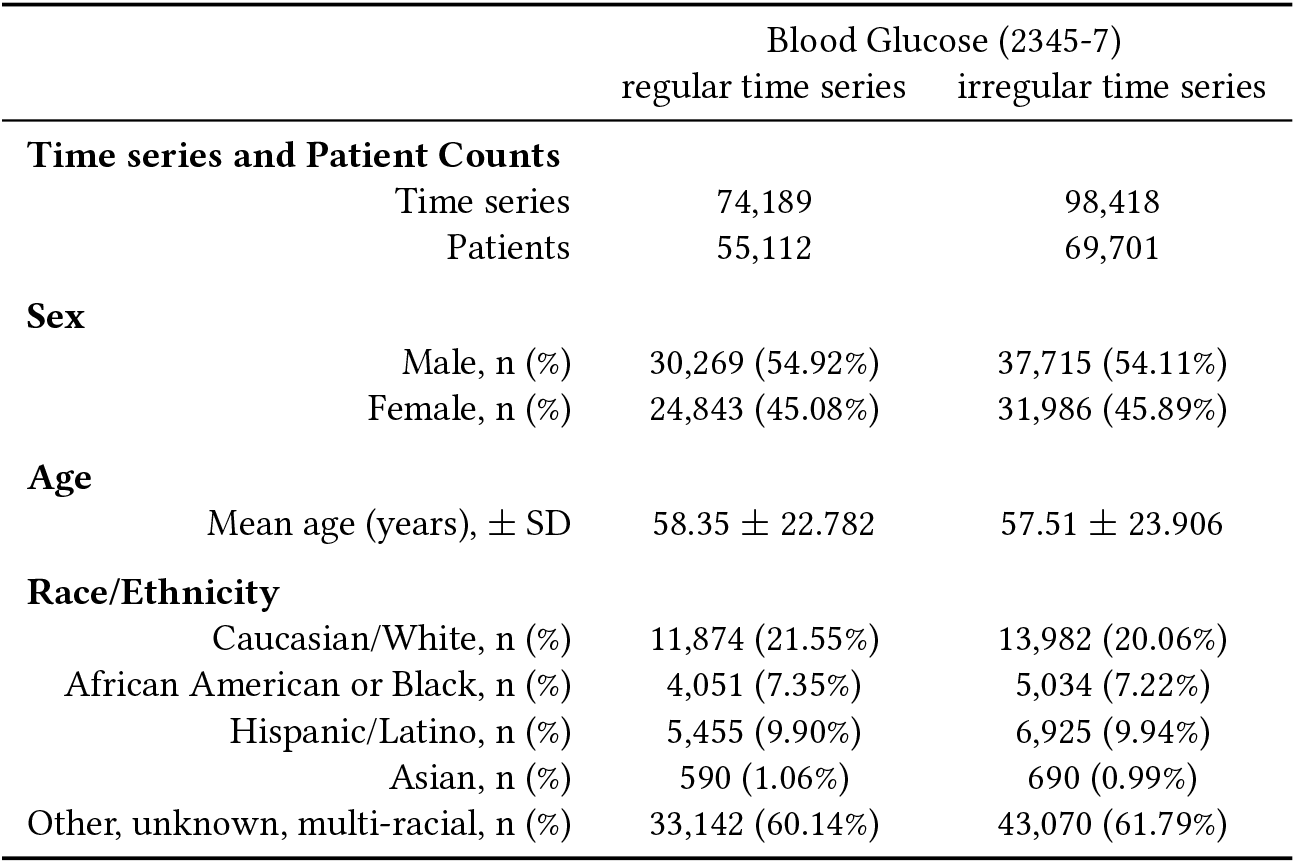
Post quality control filtering demographics for regularly and irregularly sampled blood glucose time series (days=10)

|  |  | Blood Glucose (2345-7) |  |
| --- | --- | --- | --- |
|  |  | regular time series | irregular time series |
| **Time series and Patient Counts** |  |  |  |
|  | Time series | 74,189 | 98,418 |
|  | Patients | 55,112 | 69,701 |
| **Sex** |  |  |  |
|  | Male, n (%) | 30,269 (54.92%) | 37,715 (54.11%) |
|  | Female, n (%) | 24,843 (45.08%) | 31,986 (45.89%) |
| **Age** |  |  |  |
| | Mean age (years), ± SD | 58.35 ± 22.782 | 57.51 ± 23.906 |
| **Race/Ethnicity** |  |  |  |
|  | Caucasian/White, n (%) | 11,874 (21.55%) | 13,982 (20.06%) |
|  | African American or Black, n (%) | 4,051 (7.35%) | 5,034 (7.22%) |
|  | Hispanic/Latino, n (%) | 5,455 (9.90%) | 6,925 (9.94%) |
|  | Asian, n (%) | 590 (1.06%) | 690 (0.99%) |
|  | Other, unknown, multi-racial, n (%) | 33,142 (60.14%) | 43,070 (61.79%) |

**Figure 5:**
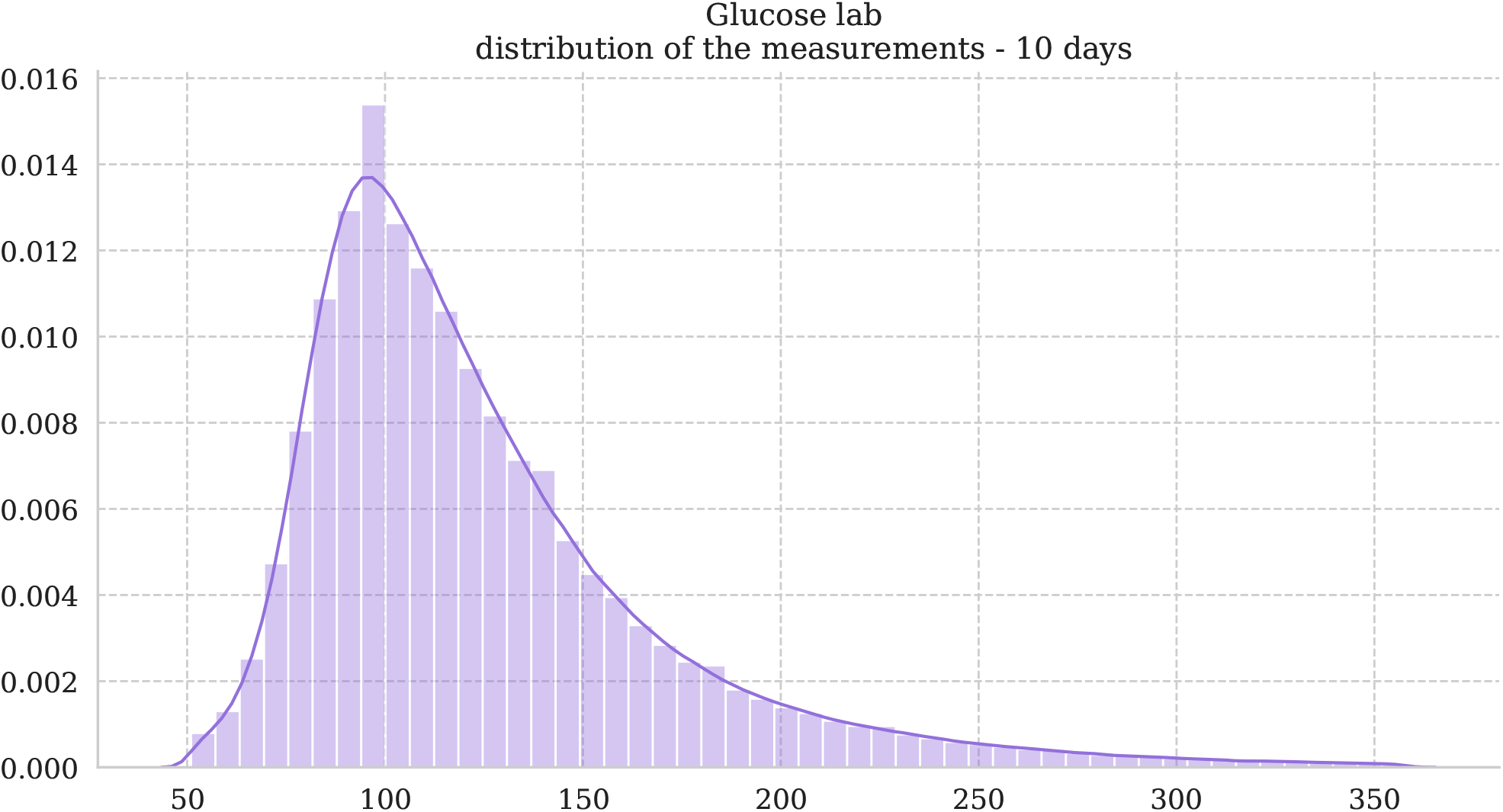
Distribution of glucose lab measurements post-quality control for irregular time series.

**Figure 6:**
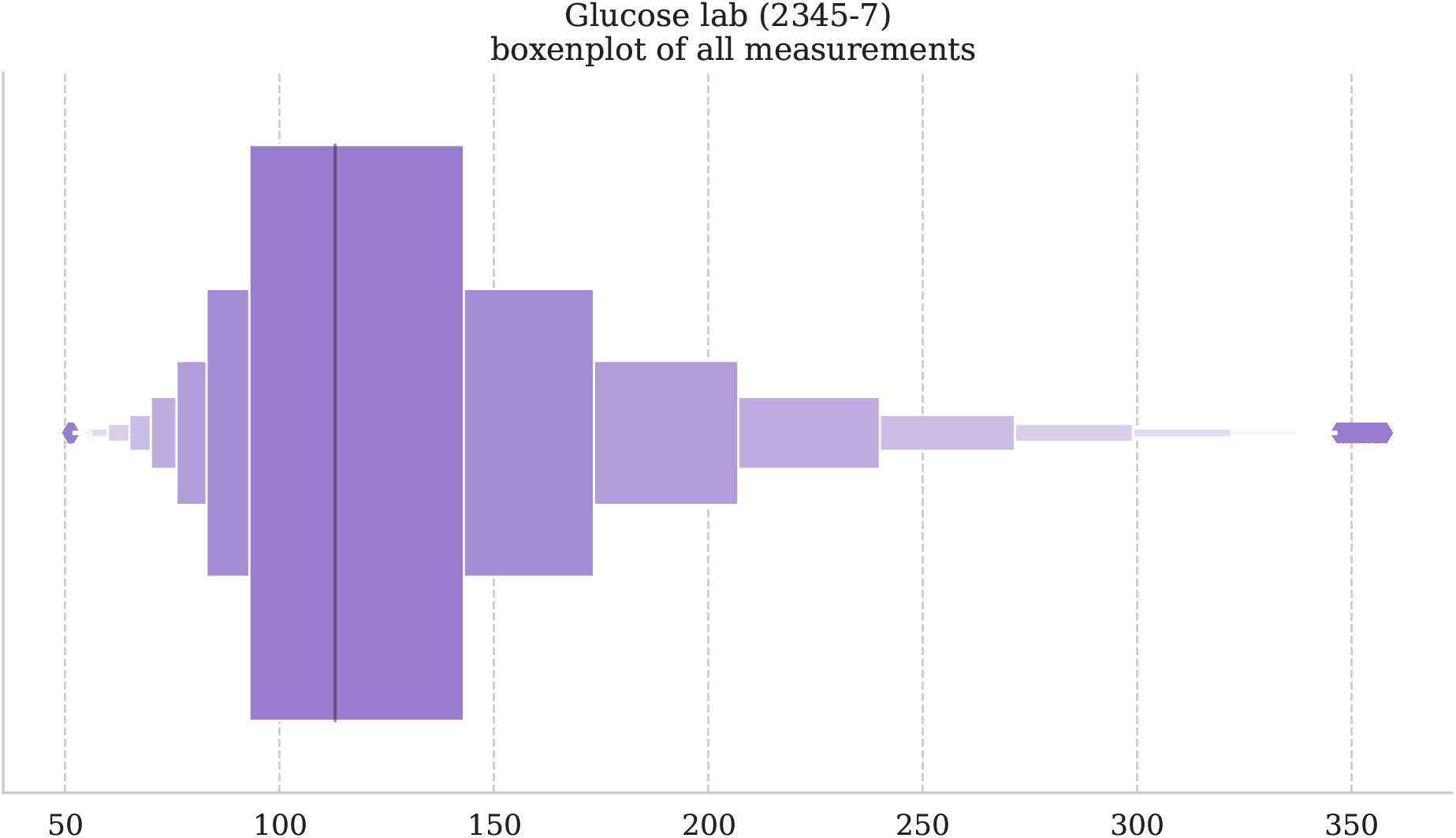
Boxenplot of glucose lab measurements post-quality control for irregular time series to visualize percentiles and potential outliers.

**Figure 7:**
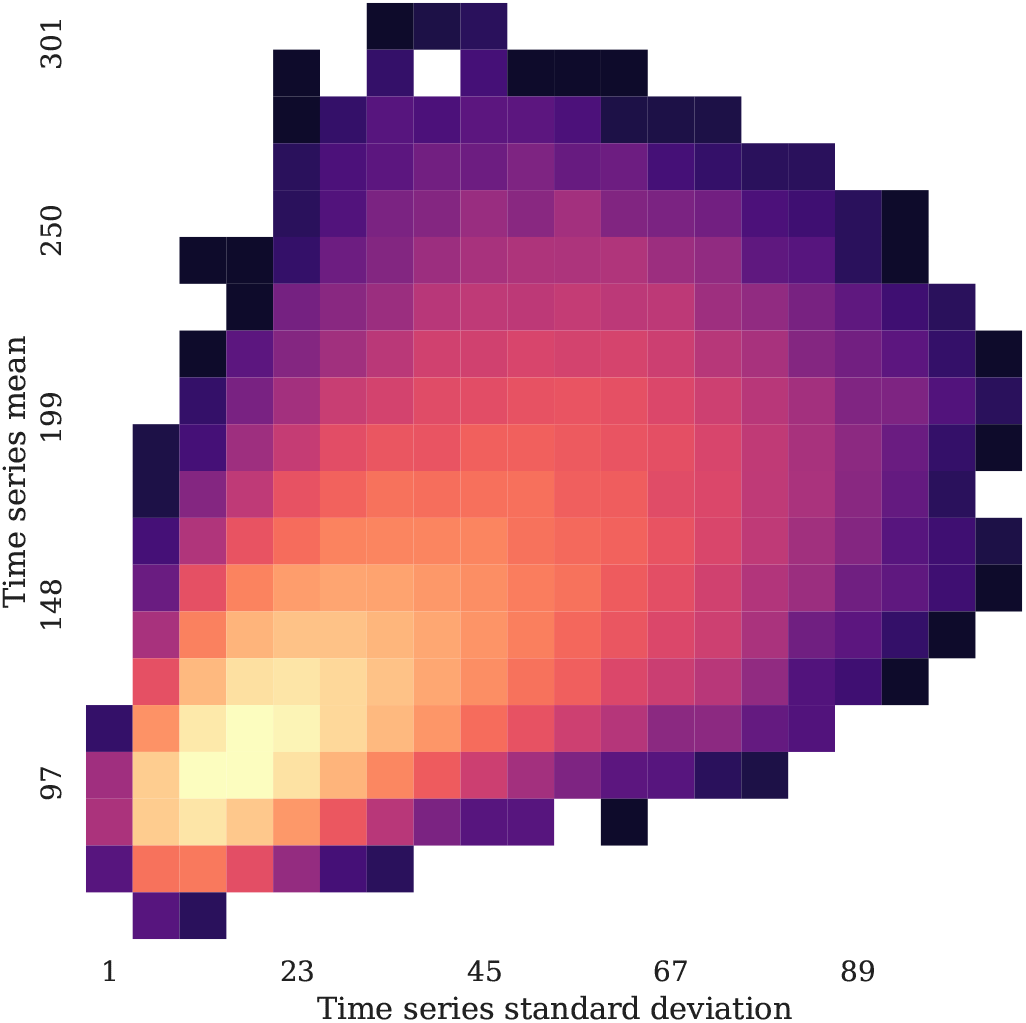
Density heatmap of regular glucose lab measurements time series represented by their standard deviations and mean.

#### 2.1.3 Drug exposure information

The drug exposure information were used in the conditional WGAN as auxiliary information, to be able to later simulate time series based on real or hand-picked drug exposure conditional vectors. For each time series, we collected the drugs at the ingredient level (i.e., referenced by RxNorm) that had a drug era overlap with the measurements.

Drug eras were defined at the ingredient level using the definition of the OMOP CDM: they are extrapolated from drug exposures with a persistence window of 30 days, meaning that prescriptions with a gap lesser or equal to 30 days belong to the same drug era (Figure 9).

**Figure 8:**
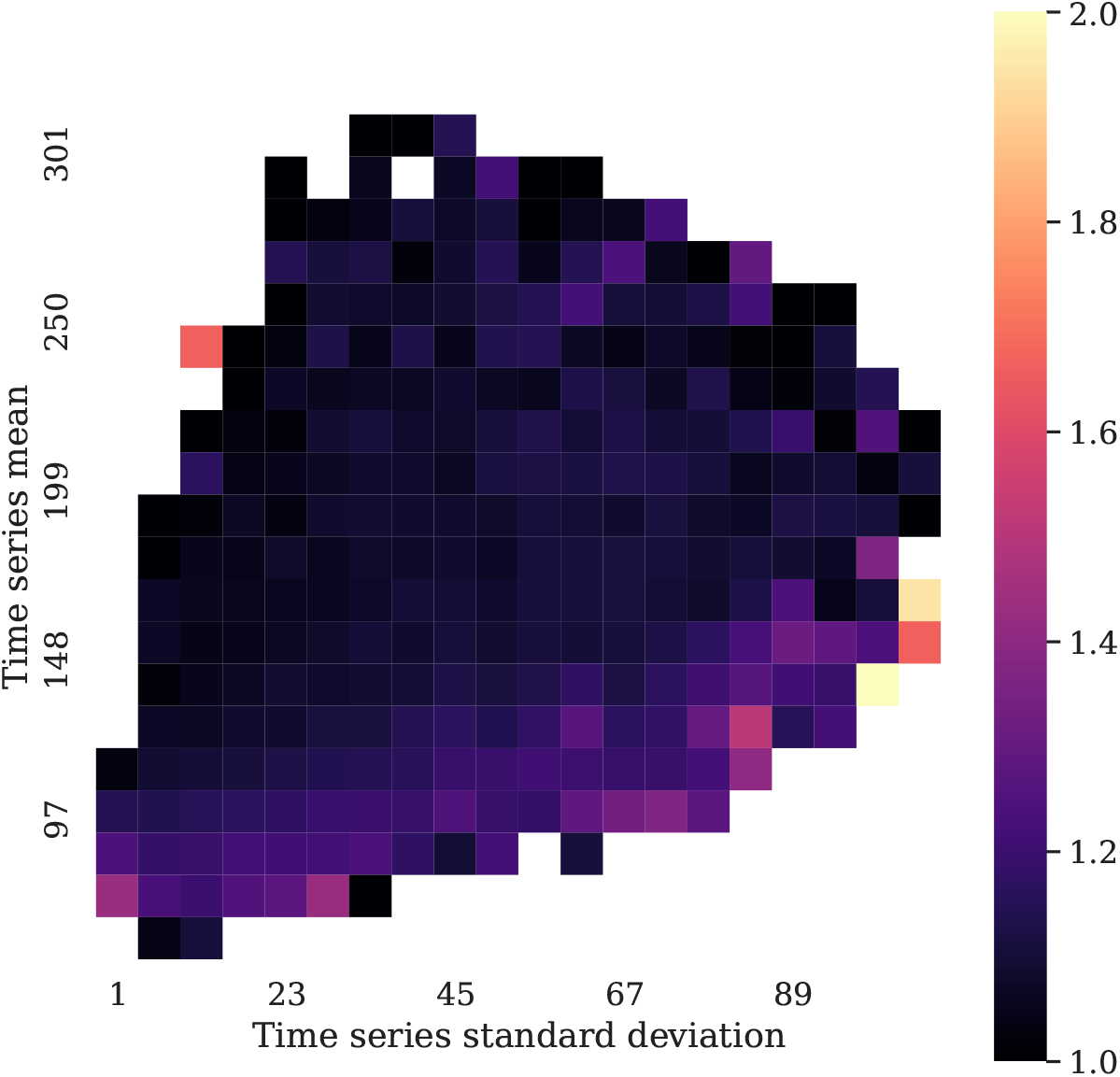
Time intervals heatmap of regular glucose lab measurements time series represented by their standard deviations and mean.

**Figure 9:**
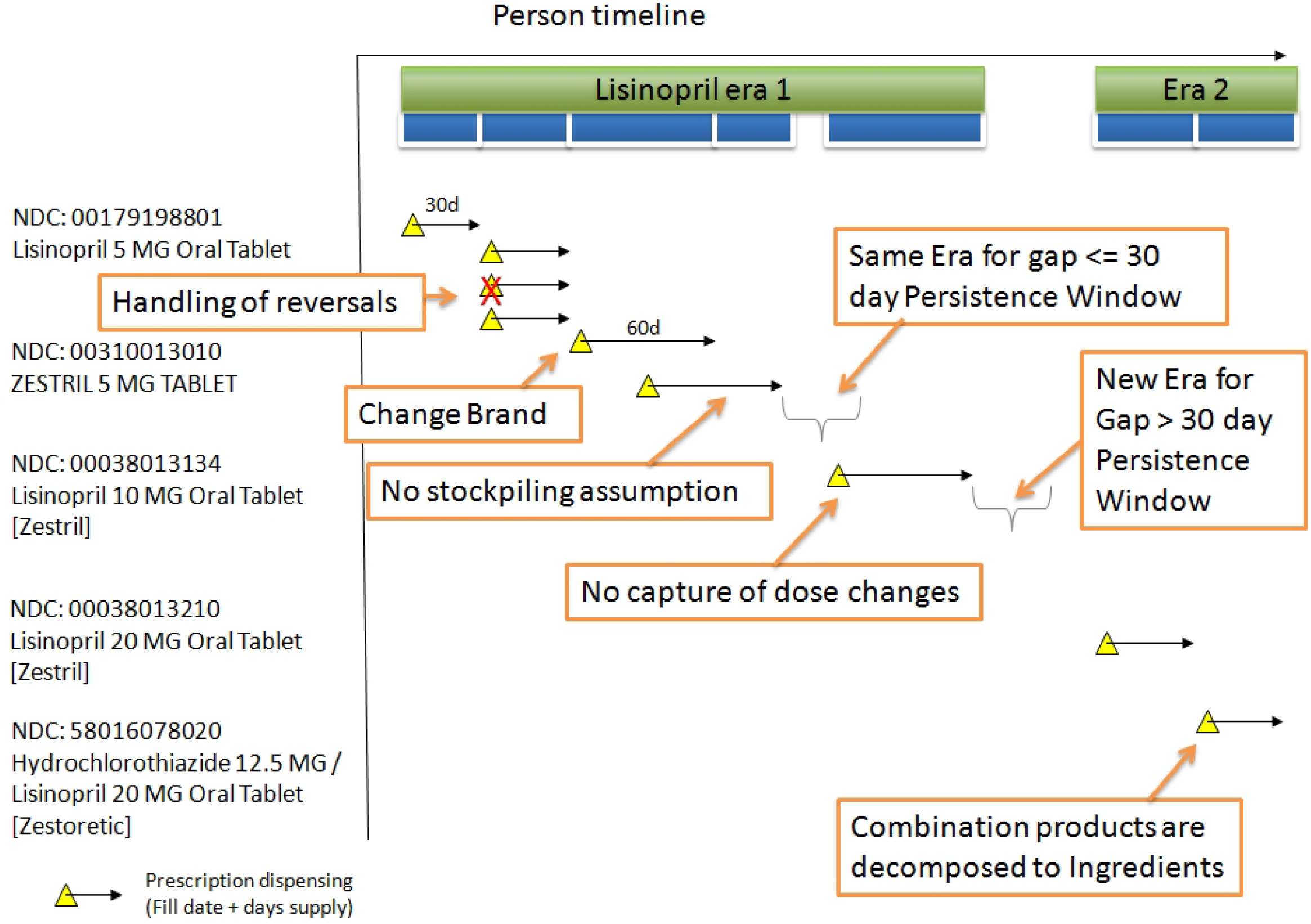
Construction of drug eras in the OMOP CDM

The Anatomical Therapeutic Chemical (ATC) classification system [1] is a hierarchical terminology controlled by the World Health Organization Collaborating Centre for Drug Statistics Methodology (WHOCC) to group drugs (Table 5). We mapped these RxNorm codes to their ATC counterparts. This RxNorm to ATC mapping is one-to-many, but it presents the advantage of enabling hierarchical grouping.

**Table 5:**
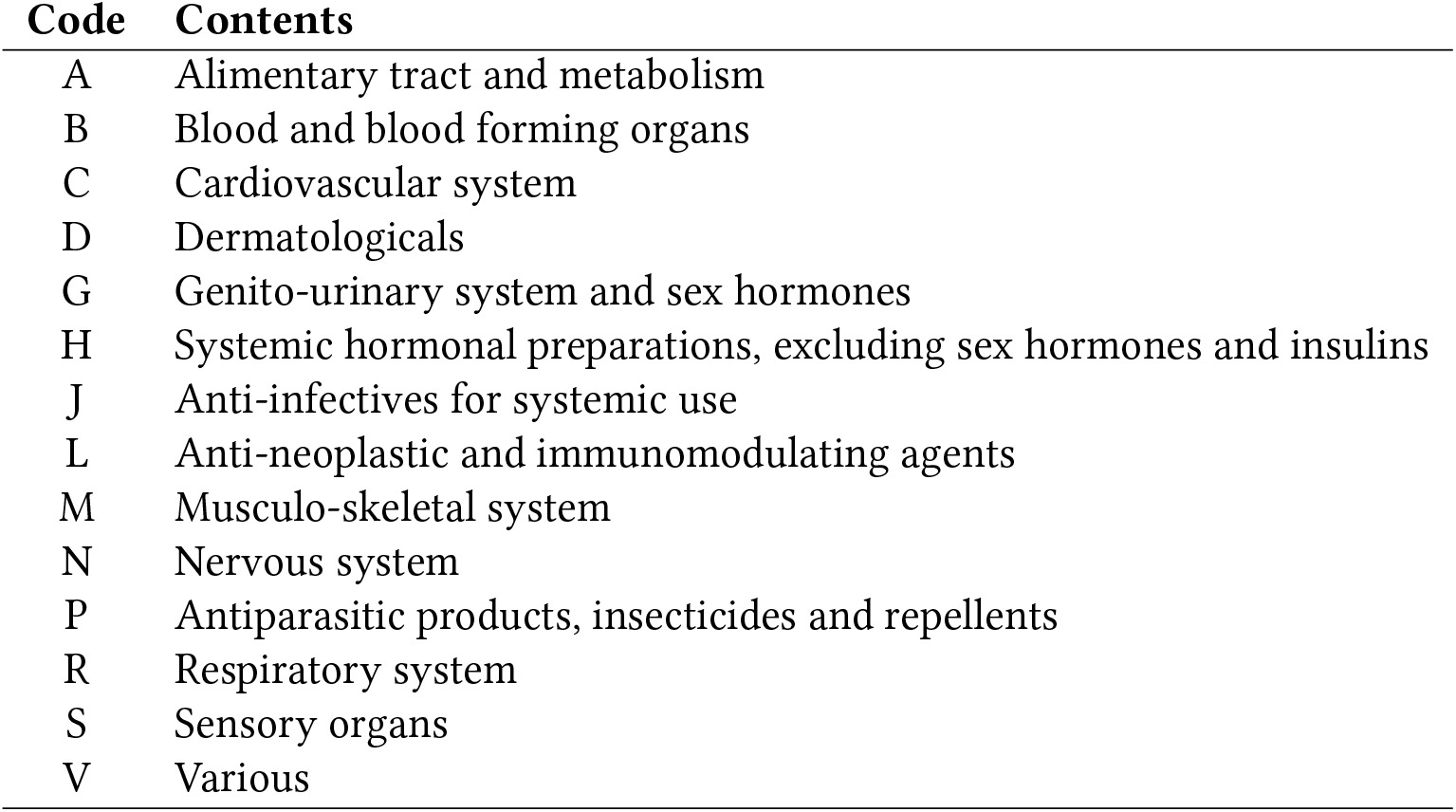
First level of the Anatomical Therapeutic Chemical (ATC) classification system

| Code | Contents |
| --- | --- |
| A | Alimentary tract and metabolism |
| B | Blood and blood forming organs |
| C | Cardiovascular system |
| D | Dermatologicals |
| G | Genito-urinary system and sex hormones |
| H | Systemic hormonal preparations, excluding sex hormones and insulins |
| J | Anti-infectives for systemic use |
| L | Anti-neoplastic and immunomodulating agents |
| M | Musculo-skeletal system |
| N | Nervous system |
| P | Antiparasitic products, insecticides and repellents |
| R | Respiratory system |
| S | Sensory organs |
| V | Various |

ATC categories represent different granularity levels: therapeutic subgroups (second level), therapeutic/pharmacological subgroup (third level), and chemical/therapeutic/pharmacological subgroup (fourth level), a granularity that resembles the most drug classes. We focused on the third (ATC-3), fourth (ATC-4) and fifth level (ATC-5, ingredient level similar to RxNorm).

For each of the four representations, we performed a two-sample Kolmogorov-Smirnov test to assess how different the distributions of the means of the laboratory test time series (LTTS) were between the exposed and non-exposed groups. We ranked them by p-value, adjusted for multiple hypothesis testing since we re-used samples between the different tests, and KS statistics. The drug exposure vector was then built using the 5 or 10 most significant drugs for each of the four representations: ATC-3 (Table 6), ATC-4 (Table 7), ATC-5 (Table 8) and RxNorm (Table 9).

**Table 6:**
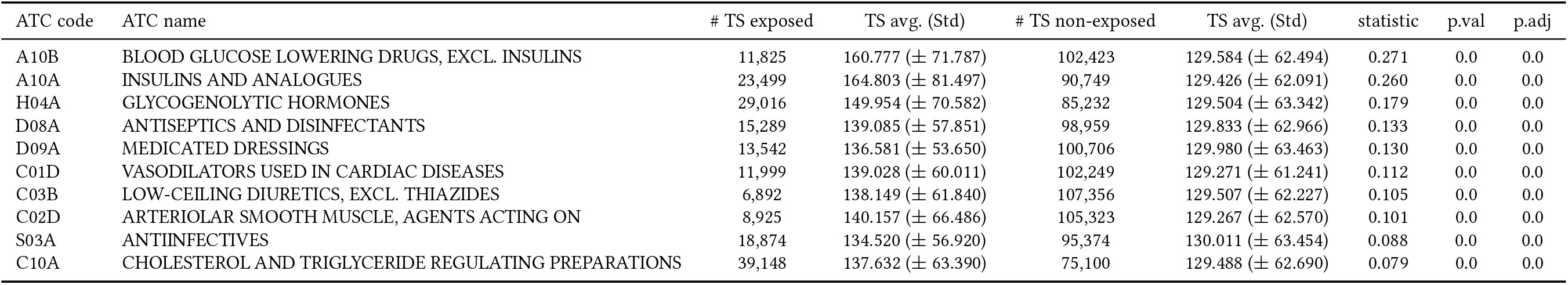
ATC level 3 abstraction of drug exposures with the most significantly different glucose lab (2345-7) time series (TS) ranked by KS statistics.

**Table 7:**
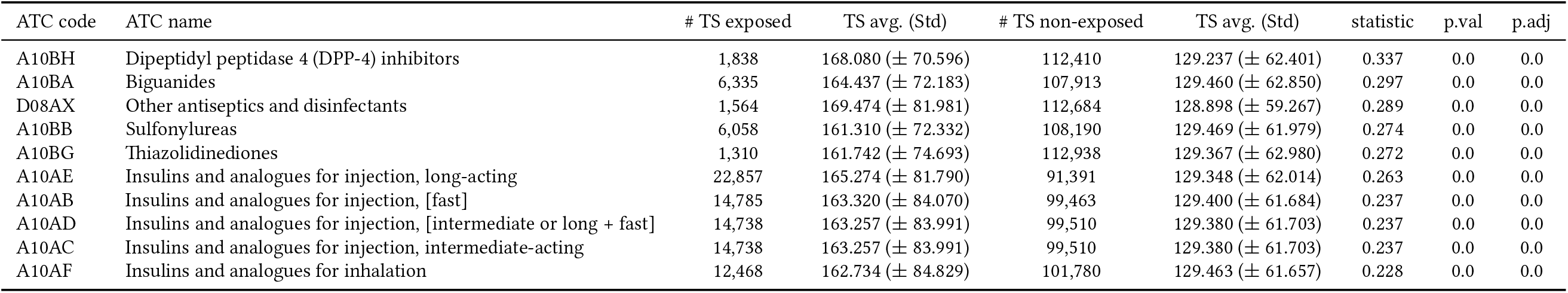
ATC level 4 abstraction of drug exposures with the most significantly different glucose lab (2345-7) time series (TS) ranked by KS statistics.

**Table 8:**
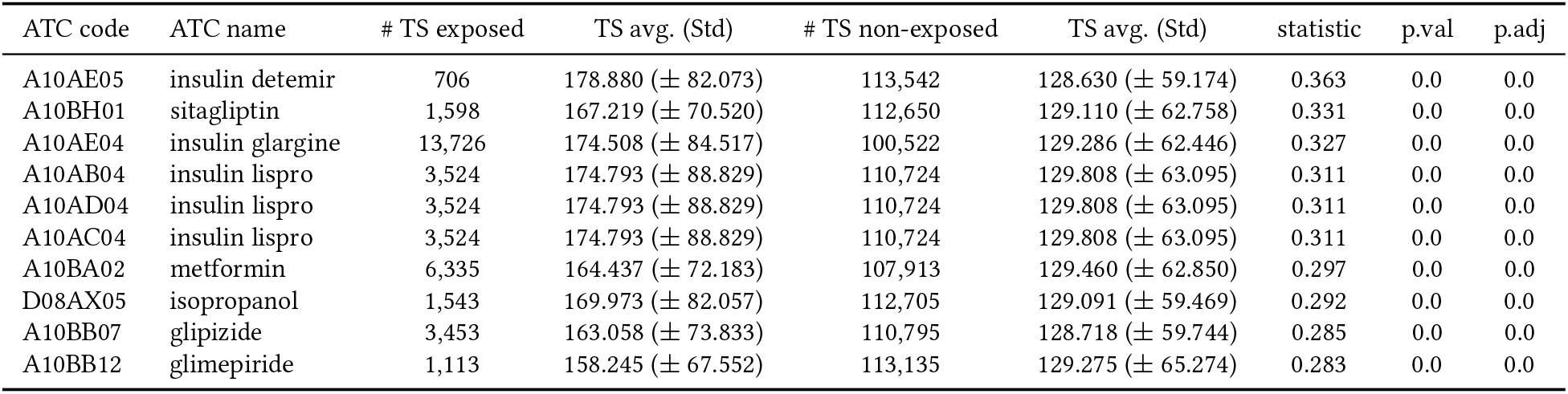
ATC level 5 abstraction of drug exposures with the most significantly different glucose lab (2345-7) time series (TS) ranked by KS statistics.

**Table 9:**
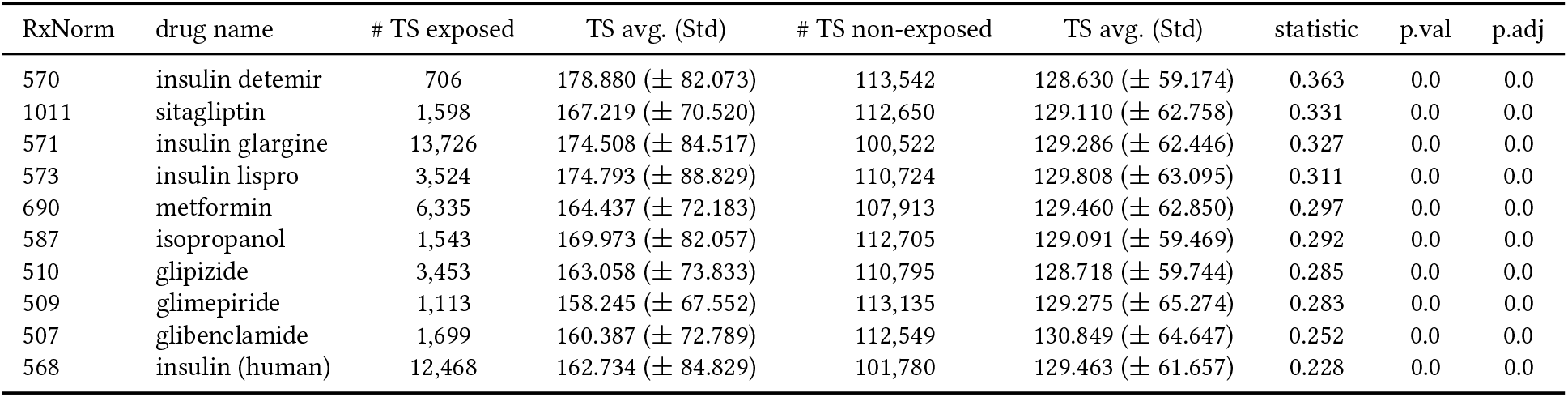
RxNorm mapping of drug exposures with the most significantly di.erent glucose lab (2345-7) time series (TS) ranked by KS statistics.

In addition, we characterized the drug exposure relationship with the laboratory test time series by representing the density heatmaps of time series exposed to the top-10 drugs according to the KS test, for each drug representation (Figures 10, 11, 12 and 13).

**Figure 10:**
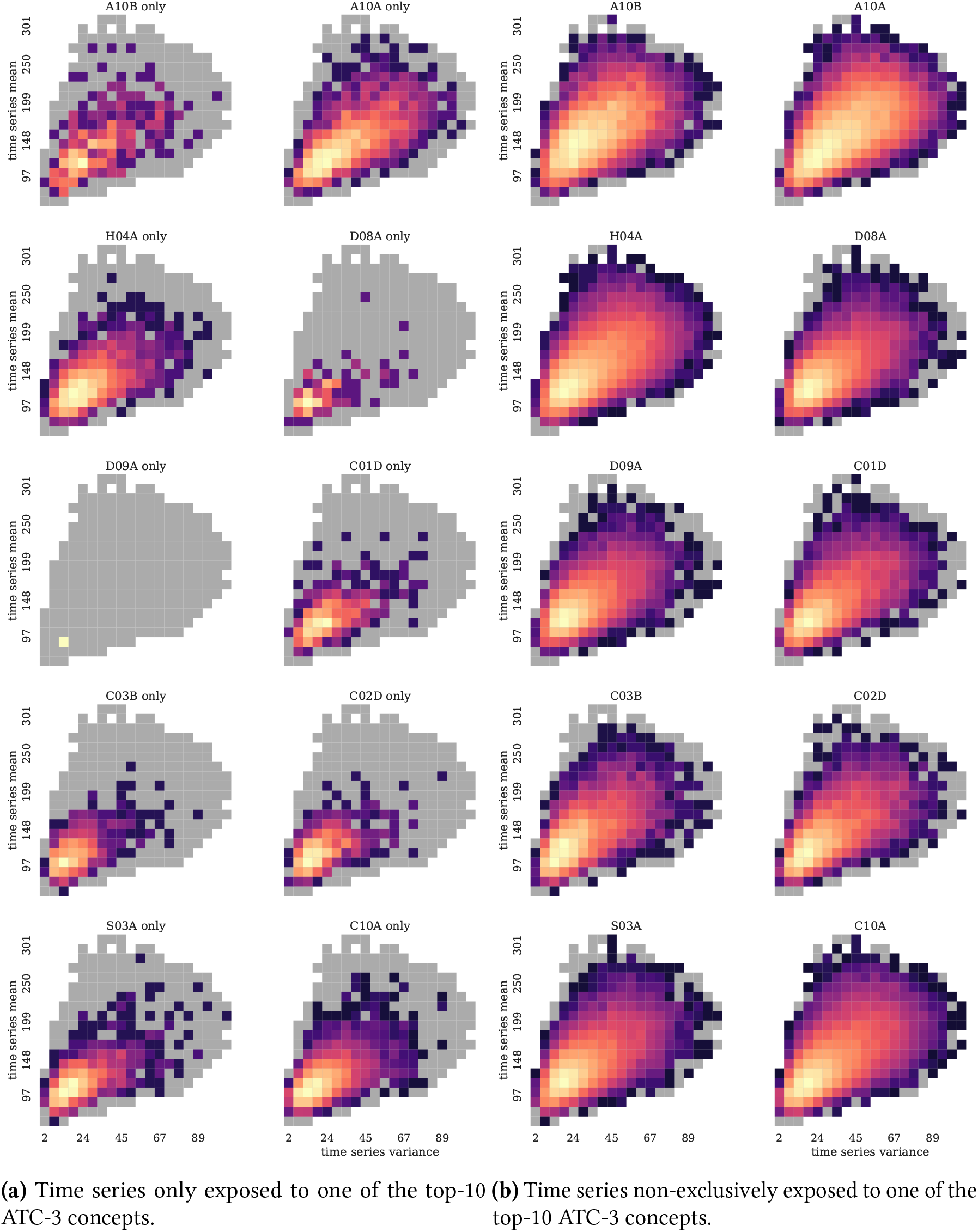
Density heatmap of irregular glucose lab time series exposed to the top-10 ATC-3 concepts ranked by KS statistics.

**Figure 11:**
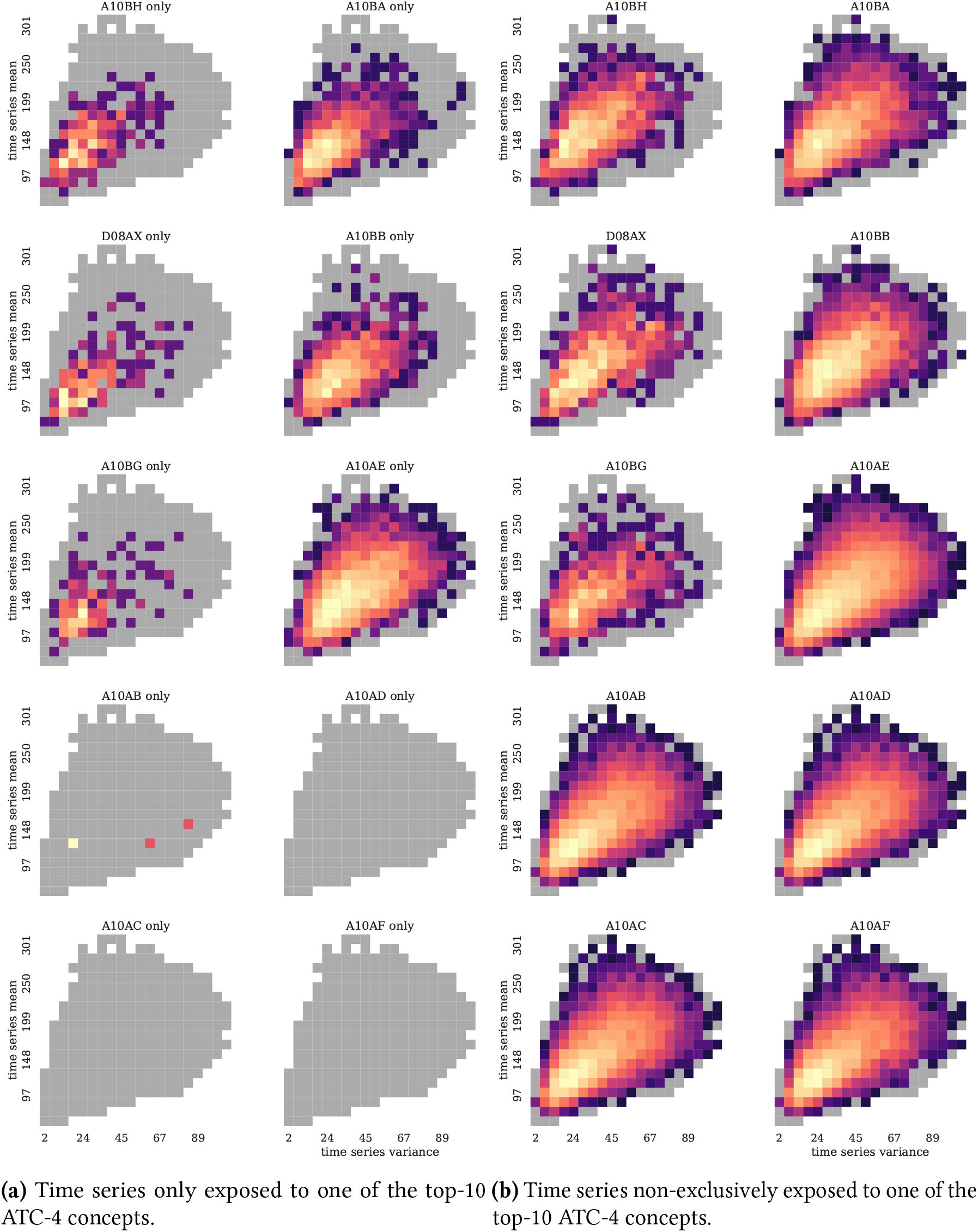
Density heatmap of irregular glucose lab time series exposed to the top-10 ATC-4 concepts ranked by KS statistics.

**Figure 12:**
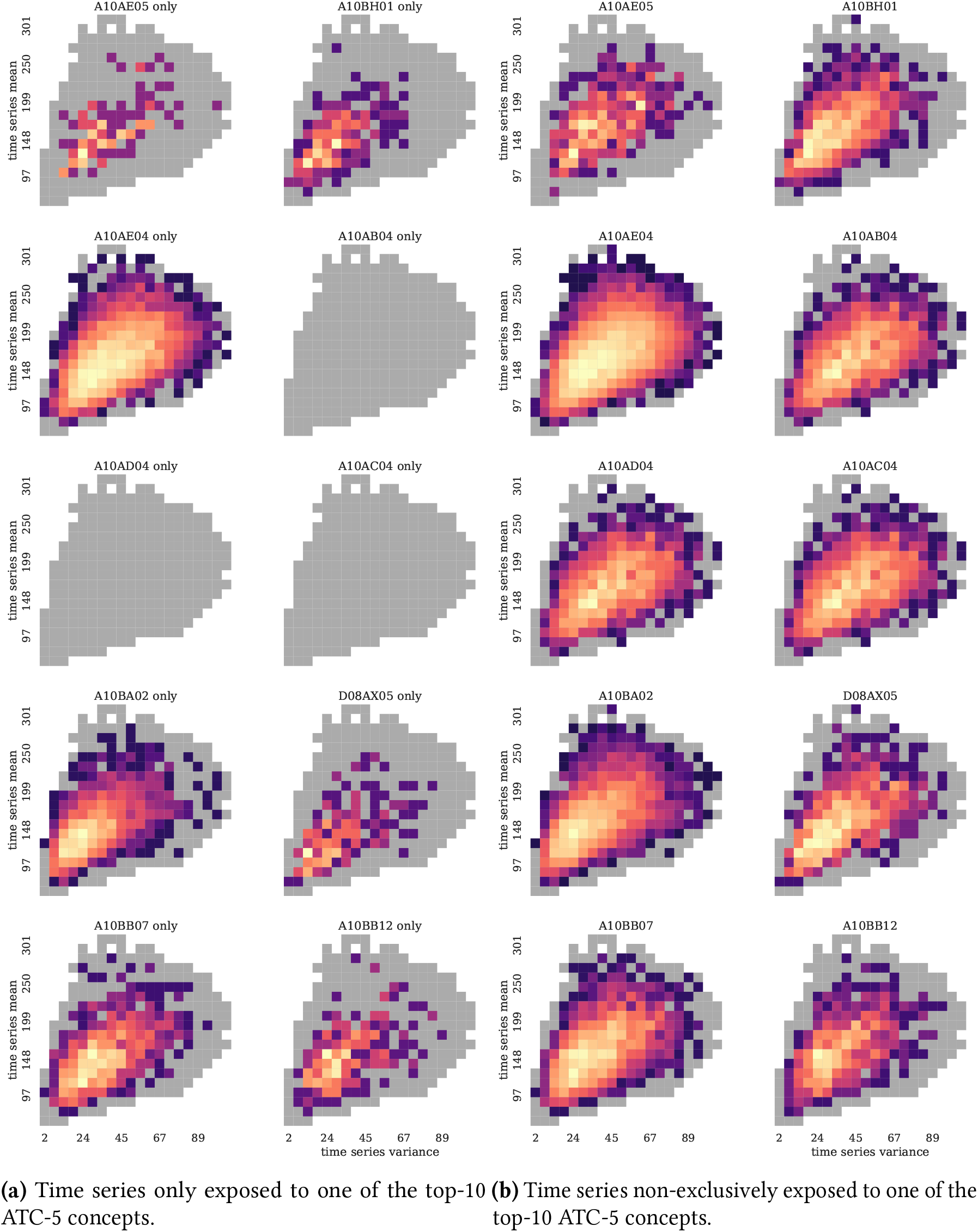
Density heatmap of irregular glucose lab time series exposed to the top-10 ATC-5 concepts ranked by KS statistics.

**Figure 13:**
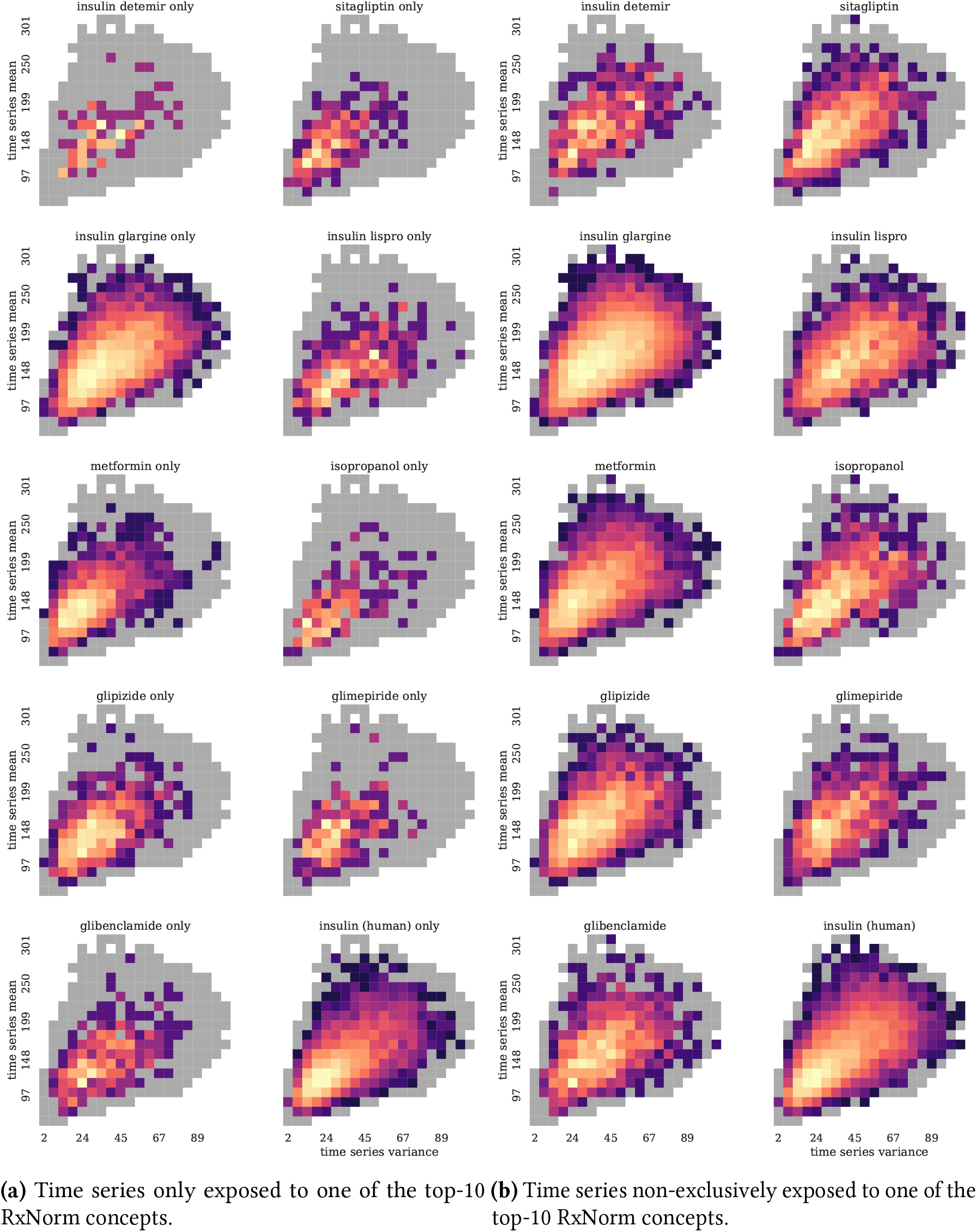
Density heatmap of irregular glucose time series exposed to the top-10 RxNorm concepts ranked by KS statistics.

It is important to note that these heatmaps show that some drug concepts are never occurring by themselves and always in combination with another top-10 drug concept. Moreover, the center of gravity of these distributions is usually higher in mean and standard deviation than the time series exposed to none of the top-10 drug concepts.

### 2.2 Deep learning models

#### 2.2.1 Deep learning experimental pipeline

Deep learning models require a more extensive hyper-parameter tuning. While their flexibility and capacity make them extremely powerful universal non-linear approximators, it comes at the cost of having to explore very large hyper-parameter spaces. We designed an experimental pipeline that balances breadth of hyper-parameters exploration, and depth of model exploration.

We split our dataset with 90/10 for training and testing. These two sub dataset were identical for the generative models and the forecasting models described in the *Supplementary Material*. For each deep learning model, we first ran a large amount of experiments with a limited amount of epochs (i.e., 50 epochs or less), determined model by model to be sufficient to observe a plateau in the training and validation loss. for these *general tuning* experiments, the loss was computed on the training set and validation set, and the testing set is held out until the very end of the process.

Following the *general tuning*, we selected the 10 models that have the lowest validation loss averaged over the second half of the epochs. For the generative models, we used the Fréchet distance to compare hyperparameter sets, estimate 10 times between randomly generated samples and the real training dataset. These models were then re-run for 100 epochs, 10 times each with different random seeds to compute more robust estimates of the performances of these models: this is the *Jne tuning* step. Based on these estimates, we selected the model (i.e., hyper parameters set) and the epoch that had the best validation loss averaged over the 10 separate runs. Generative models went through a double evaluation described in the *Evaluation metrics* section.

Concerning the forecasting models, during the *testing* step we ran the selected model for the number of epochs determined with *Jne tuning* on the whole training set and computed MSE and MAE on the test set with the final model. This step was repeated 10 times to account for the stochastic nature of neural network training and produce estimates of the MSE and MAE.

#### 2.2.2 WGAN and conditional WGAN: model specifications

The Wasserstein GAN algorithm implemented followed the recommendations of the original paper on WGAN with gradient penalty [26], including the use of layer normalization in the critic instead of batch normalization. The difference between gradient and Lipschitz penalty [44] is simply that the latter takes the maximum of zero or the gradient penalty, ensuring in effect that the gradient penalty is always positive or null.

The conditional architecture was designed by transposing the conditional GAN paper [41] to the WGAN framework: the auxiliary information were concatenated with the random latent vector at the input of the generator, to generate the time series. It was then concatenated again to the produced synthetic time series, and to the real time series at the input of the critic, so that the critic also computes the estimated Wasserstein-1 distance taking into account the auxiliary information – particularly important for the generator’s training. Finally, we used RMSProp for the optimization of the WGANs, following the recommendations of Gulrajani et al, and Petzka et al. [26, 44]. The objective functions can be found in Supplementary Materials.

### 2.3 Evaluation metrics

Evaluating implicit models is a hard task. In computer vision for instance, where human inspection can be used as a sanity check, a dozen of different evaluation metrics have been proposed to compare real and synthetic data and quantify how close the stochastically generated samples are from the training data.[9] With EHR data, we do not have the luxury of visual inspection or highly engineered computer vision networks, and expert evaluation by physician is both time-consuming and costly, and arguably not precise enough to catch subtle differences in very large datasets. More importantly, while computer vision benefits from standard datasets such as MNIST [36], CIFAR-10[34], CelebA[69], or ImageNet [15], biomedical data sciences do not have standard medical datasets, mainly due to privacy and regulations, although MIMIC-III is increasingly regarded as such.[30]

Regardless, in spite of the absence of consensus regarding standard datasets, we must be able to design quantitative evaluation metrics for implicit generative models of EHR data in a single-institution setting to begin with.

#### 2.3.1 Intrinsic evaluations

In computer vision, a wide variety of evaluation methods have been proposed to evaluate the data fidelity of synthetic images generated by GANs. Among them, an approach has been gaining momentum: inception distances. Inception distance metrics rely on the Inception Network, a heavily engineered convolutional neural network (CNN) designed to perform well on image labeling for ImageNet [59]. The intuition behind inception distances is that the weights of the penultimate layer of a deep neural network able to successfully classify images must pick up features that are high level enough to mimic the way the human visual cortex would work. Therefore, Heusel et al.[27] proposed the Féchet Inception Distance (FID) by using the distribution of weights of real and synthetic images flowed through a trained Inception network.

Let *p*_*w*_(.) be the probability of observing real data, and *p*(.) the probability of of generating model data.

The equality *p*(.) = *p*_*w*_(.) holds except for a non-measurable set if an only if: 

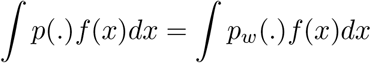

for a basis *f* (.) spanning the function space in which *p*(.) and *p*_*w*_(.) live. this function *f* (.) is replaced by the penultimate layer of an inception network trained on ImageNet and the resulting distributions of weights are approximated by multidimensional Gaussian to get the first two moments: mean and covariance. Therefore, they used the Fréchet distance [21] or Wassertein-2 distance [62] to define the Fréchet Inception Distance (FID) *d*(.,.) between the Gaussian (*m, C*) obtained from *p*(.) and the Gaussian (*m*_*w*_, *C*_*w*_) obtained from *p*_*w*_(.) given by: 

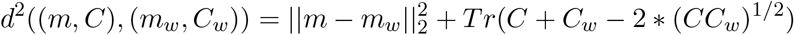

This quantitative evaluation metric for implicit generative models has shown good performance, and it has been translated to biomedical sciences for de novo drug design by Preuer et al. [45] who designed an inception network called ChemblNet, a network trained to predict bioactivities of about 1,300 assays from ChEMBL[8]. We used this Fréchet distance to evaluate the distance between the real and synthetic data generated by the GAN models we have trained, and called it FID by analogy with the FID in computer vision, although no network played the role of an inception network providing abstracted representations.

#### 2.3.2 Extrinsic evaluations

In addition of the intrinsic metrics described in the previous paragraph, we used a train on synthetic test on real (TSTR) approach, a method proposed by Esteban et al. [19], to compare the real and synthetic datasets in a supervised learning task and evaluate how well the synthetic data generated can retain the information needed for the forecasting task, and how well it generalizes.

Other task-based predictions have been described by Razavian et al.[46, 48] and Che et al.[10] using laboratory test time series for medical outcome prediction, and could be used as alternative extrinsic evaluations of the synthetic data, compared to the real data. However, these tasks are all classification tasks, while the forecasting task we describe in the *Supplementary Material* is a regression, providing a finer grained evaluation of the models and their impacts.

### 2.4 Applications of conditional WGANs: drug effect simulation in laboratory test time series

The two main directions of these experiments are: 1/ the simulation of lab test time series by manually selecting conditional drug exposure vectors, and investigating the interpolation power of these model to infer the behavior of time series with conditional information not seen during training; 2/ the data augmentation task, where real datasets are augmented with synthetic data to improve locally forecasting performances using the regressors described in the *Supplementary Material*.

#### 2.4.1 Inference power evaluation

In order to evaluate how well the conditional WGANs trained in the previous Aim 2 can infer samples from conditional class unseen at training, we designed the following experiments: we removed from the training dataset glucose time series exposed to a unique drug, trained the GAN from scratch on this new dataset, and then generated samples with single drug exposures to compare them to real single-drug time series, and single-drug time series from the Aim 2 WGAN model exposed to them at training. This experiment relies on the interpolation power of the latent space in GANs. The two hypotheses tested in this evaluation are that conditional GAN can infer the behavior of samples from other classes seen at training (i.e., even if the GAN has never seen drug A alone, it has seen drug A in combination with other drug exposures), and that the latent space can continuously interpolate these inferred samples. It means that we could for a given drug of exposure generate all the states between its two binary states, 0 and 1, and get continuously closer to samples exposed to the drug.

In order to evaluate the simulation of these time series, we used the Frechet Inception Distance (FID) introduced in the previous section as a distance metric.

#### 2.4.2 Data Augmentation by conditional generation

After analyzing the simulation properties of conditional WGANs, we investigated their usage for data augmentation. If these models can infer unseen classes, or unseen combinations of classes, they could inflate specific sub-groups of samples in training sets to improve the performances of the model at testing. In this second experiment, we first augmented time series with unique drug exposures and evaluated the performances of the MLP forecasting model from Aim 1 with these new training datasets.

Then, we targeted specific drug combinations that have low frequency in the training set, and on which forecasting models yield high error on the testing set. These are the most interesting candidates for data augmentation, as they could benefit from a high count in the training set to decrease their high testing errors.

## 3 Results

### 3.1 Regular LTTS generation with WGAN-GP

#### 3.1.1 Hyperparameter tuning

The WGAN hyperparameter tuning was extensive, with more than 7,000 hyperparameter combinations evaluated (Table 10).

**Table 10:** Hyper-parameters for WGAN general tuning on regular glucose lab time series.

| Parameters | Round 1 | Round 2 | Round 3 |
| --- | --- | --- | --- |
| Layers width | 128 | 64, 128 | 128 |
| Number of layers | 2 | 2, 3 | 2, 3, 5 |
| leakyReLU | 1e-1, 1e-3 | 1e-1, 1e-2, 1e-3 | 1e-1, 1e-2, 1e-3 |
| Dropout | 0.25, 0.50 | 0.25, 0.50 | 0.0, 0.25 |
| Batch Size | 32 | 64, 128 | 32, 64, 128 |
| Critic Update cycle | 10, 5 | 10, 5 | 10, 5 |
| Critic learning rate | 1e-4, 5e-5 | 5e-5 | 1e-4, 5e-5 |
| Generator learning rate | 1e-4, 5e-5 | 5e-5 | 1e-4, 5e-5 |
| Weight decay (L2) | 1e-3, 1e-4 | 5e-3, 1e-4, 0.0 | 5e-3, 1e-4, 0.0 |
| Lambda penalty | 10, 5, 1 | 10, 5, 1 | 5, 1, 0.5, 0.1 |
| Latent dimension | 100, 10 | 100, 10 | 100 |
| Num. combinations | 384 | 1,728 | 5,184 |

We took the best performing hyperparameter combinations in terms of FID, and tested them, adding gradient penalty (GP) to the Lipschitz penalty (LP) (Table 11) used for the general tuning above.

**Table 11:**
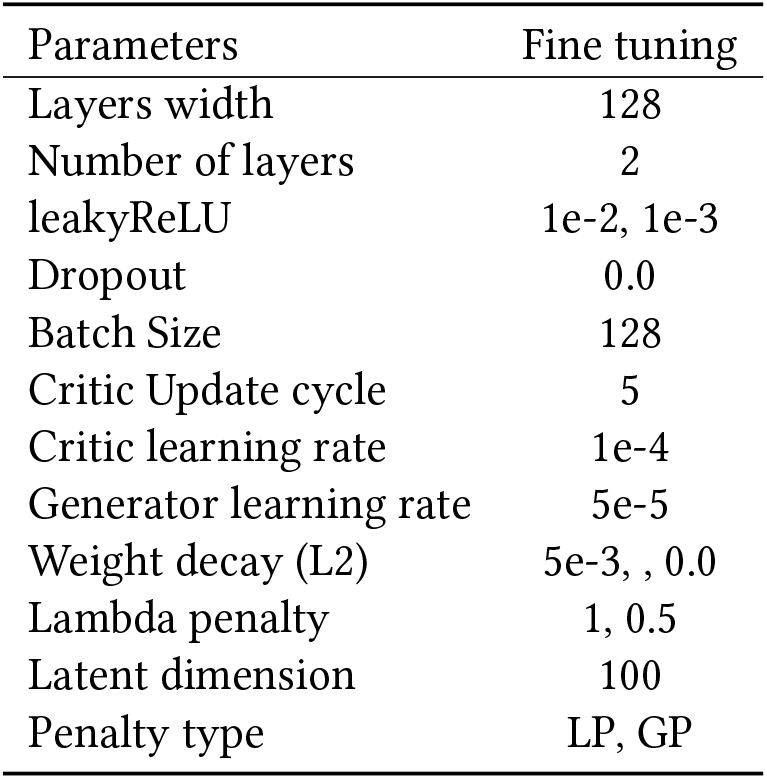
Hyper-parameters for WGAN fine tuning on regular glucose lab time series.

#### 3.1.2 Impact of sampling on FID

For each ratio of synthetic data, we generated 10 distinct datasets to get an estimate of the FID and how much it varies. The FID values and their sub-components (i.e, difference of the means and difference of the covariance matrices) did not change significantly depending on how much synthetic data was generated to compute the distance. However, the standard deviation tended to decrease as the amount of synthetic data increases, as expected (Figure 14).

**Figure 14:**
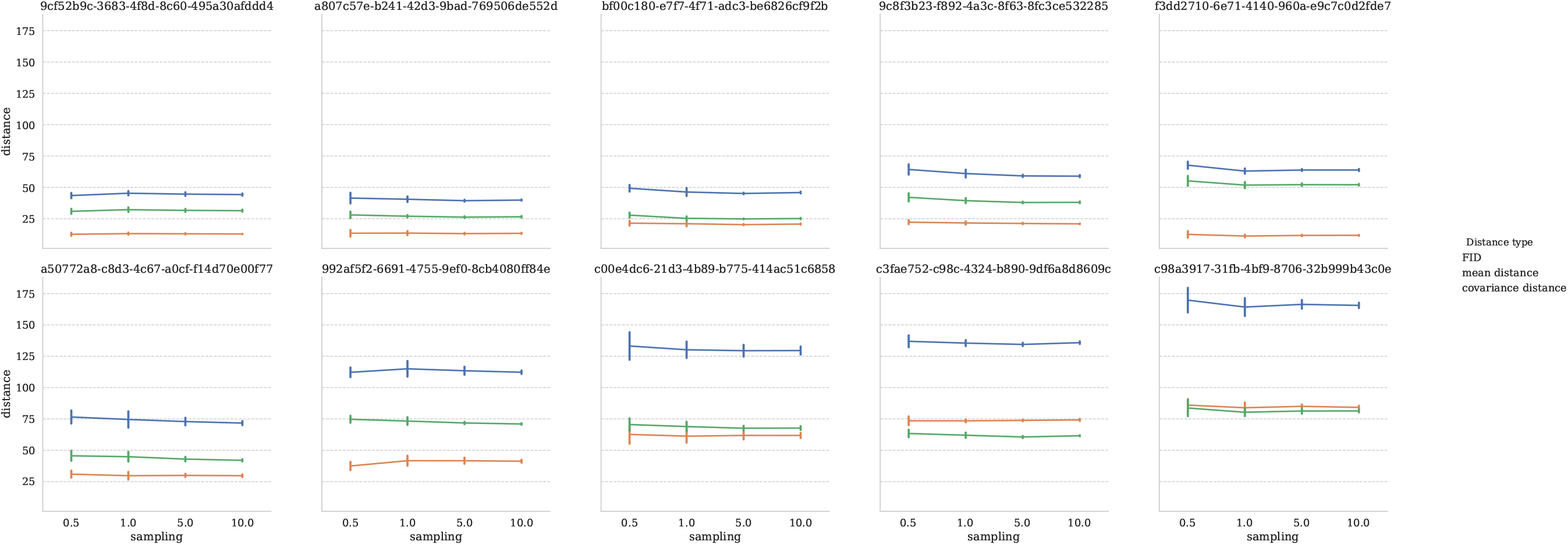
Evolution of the FID for different amounts of sampled synthetic data, expressed in ratio of real data, for WGANs on regular glucose lab time series.

#### 3.1.3 FID evolution during training

The FID dropped within the first 30 epochs or so, and then either stabilized, or showed signs of overfitting of the WGAN with an upward trend toward epoch 80 (Figure 15). For reference, the FID between the real training and the testing set was 15.22 (mean diff.: 4.85, cov. diff.: 10.37) while the FID of the top 10 GAN model spanned from 26.71 to 46.26.

**Figure 15:**
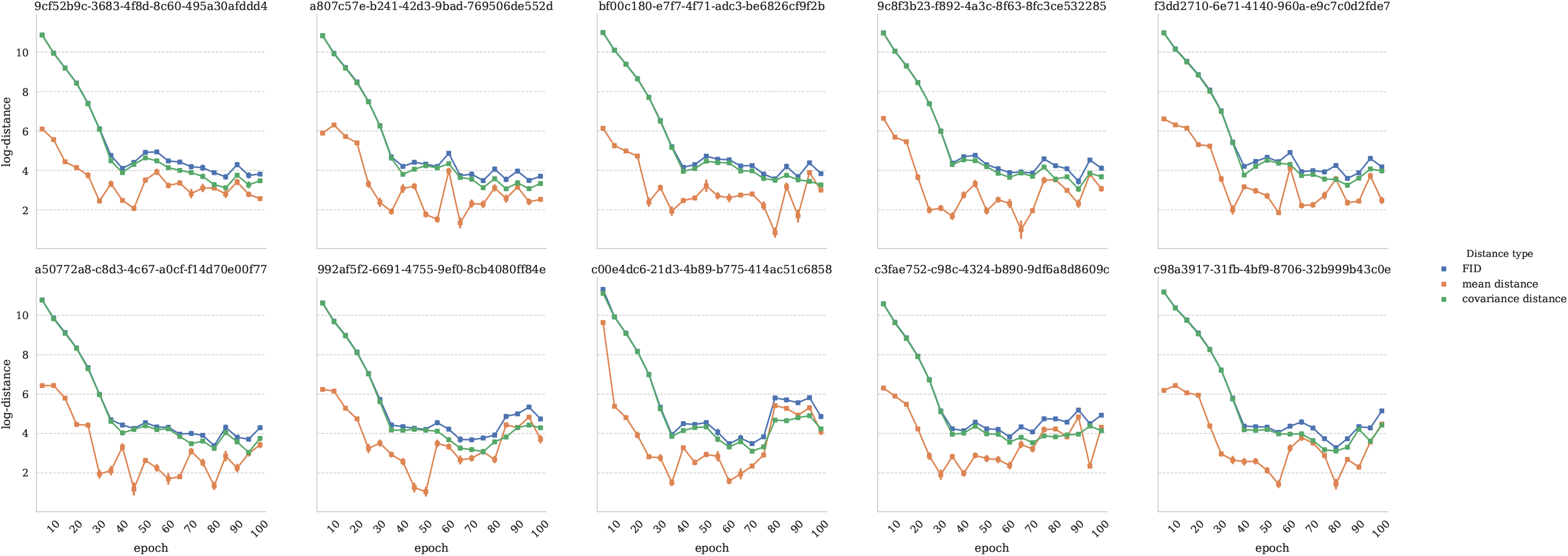
Evolution of the FID during training of WGANs on regular glucose lab time series every 5 epochs.

#### 3.1.4 TSTR with an MLP forecasting model

We evaluated the 10 best WGAN models according to their FID with the TSTR extrinsic evaluation (Table 12). We can see that the difference in the covariance matrices was more correlated to the TSTR metrics than the FID itself. The general trend is a relatively low error on forecasting synthetic samples from the same generator, a higher error on the training data that the WGAN was trained on, and the highest error on the testing set that the WGAN was never exposed to. While the MSE on the testing set was above 1,100, it is not extremely far from the error from the models trained on real regular time series (see *Supplementary Material*). The GAN model that performed the best at the TSTR task got an average MSE of 1120.84 (*±*8.553), and a lowest MSE at 1103.84. Interestingly enough, it was not the model with the lowest FID (39.88).

**Table 12:** Train on Synthetic Test on Real (TSTR) evaluation of the top-10 WGAN models selected by FID.

| Model | avg.<br>FID (±SD) | avg. mean<br>diff. (±SD) | avg. cov.<br>diff (±SD) | best<br>synth. error | avg synth.<br>error (±SD) | best real<br>training error | avg. real<br>train error (±SD) | best real<br>test error | avg. real<br>test error (±SD) |
| --- | --- | --- | --- | --- | --- | --- | --- | --- | --- |
| 9 | 26.71<br>(± 1.793) | 4.30<br>(± 1.001) | 22.42<br>(± 1.195) | 947.19 | 959.22<br>(± 7.740) | 1079.71 | 1092.15<br>(± 12.250) | 1119.00 | 1133.46<br>(± 13.846) |
| 5 | 29.55<br>(± 1.594) | 3.87<br>(± 0.686) | 25.67<br>(± 1.256) | 1019.16 | 1038.17<br>(± 10.404) | 1068.04 | 1082.67<br>(± 8.600) | 1131.89 | 1142.88<br>(± 11.224) |
| 3 | 31.42<br>(± 3.912) | 10.18<br>(± 1.895) | 21.24<br>(± 2.286) | 1250.11 | 1378.61<br>(± 65.012) | 1416.08 | 1549.92<br>(± 69.978) | 1494.29 | 1639.10<br>(± 75.955) |
| 7 | 32.60<br>(± 2.448) | 4.92<br>(± 0.597) | 27.68<br>(± 2.006) | 1165.10 | 1185.05<br>(± 15.426) | 1088.98 | 1105.20<br>(± 13.304) | 1128.52 | 1145.84<br>(± 11.975) |
| 1 | 32.60<br>(± 2.618) | 9.90<br>(± 1.601) | 22.71<br>(± 1.577) | 965.49 | 984.86<br>(± 12.326) | 1093.60 | 1113.47<br>(± 10.072) | 1129.14 | 1149.45<br>(± 10.614) |
| 2 | 35.48<br>(± 1.193) | 2.37<br>(± 0.534) | 33.11<br>(± 1.151) | 861.18 | 873.90<br>(± 8.863) | 1095.20 | 1110.45<br>(± 9.082) | 1158.04 | 1177.30<br>(± 13.636) |
| 4 | 36.39<br>(± 2.467) | 10.66<br>(± 1.171) | 25.73<br>(± 1.604) | 1026.15 | 1057.11<br>(± 23.045) | 1112.40 | 1139.37<br>(± 20.813) | 1165.91 | 1184.12<br>(± 18.020) |
| 0 | 39.35<br>(± 3.940) | 16.72<br>(± 2.942) | 22.64<br>(± 1.711) | 902.49 | 917.09<br>(± 11.450) | 1100.04 | 1111.24<br>(± 7.952) | 1146.76 | 1158.13<br>(± 8.773) |
| **6** | 39.88<br>(± 2.959) | 15.69<br>(± 1.872) | 24.19<br>(± 2.020) | **1002.33** | 1023.22<br>(± 11.621) | 1058.22 | 1074.86<br>(± 8.519) | **1103.84** | 1120.84<br>(± 8.553) |
| 8 | 46.26<br>(± 3.861) | 10.88<br>(± 1.715) | 35.38<br>(± 2.687) | 1040.69 | 1056.51<br>(± 11.263) | 1092.84 | 1105.79<br>(± 9.446) | 1131.36 | 1145.82<br>(± 10.643) |

#### 3.1.5 Density evolution through training of the best regular WGAN

By sampling every 5 epochs a synthetic dataset of the same size as the training dataset, we can visualize the coverage of the synthetic data compared to the real training data it was fitted on (Figure 16). We also computed the weighted ratio of synthetic coverage. On every tile of the density heatmap, We computed the difference of real and synthetic data, and counted hits (correct coverage), wrong (coverage of a tile not represented in the real data), and miss (absence of coverage of a tile represented in the real data) (Figure 17). This barplot summarizes quantitatively the heatmaps by epoch. We can observe that coverage increased fast and then plateaued around 30 epochs, to increase again past 70 epochs. The wrong ”modes”, synthetic time series that have unrealistic summary statistic, decreased within the first 30 epochs, and coverage improved after 60 epochs. When observing the kernel density estimation plot, we see that the mean distribution is approximated first, and then the covariance is adjusted in order to cover the real distribution better during that second stage of the training. The best coverage reached was 88.22% and was reached at epoch 95.

**Figure 16:**
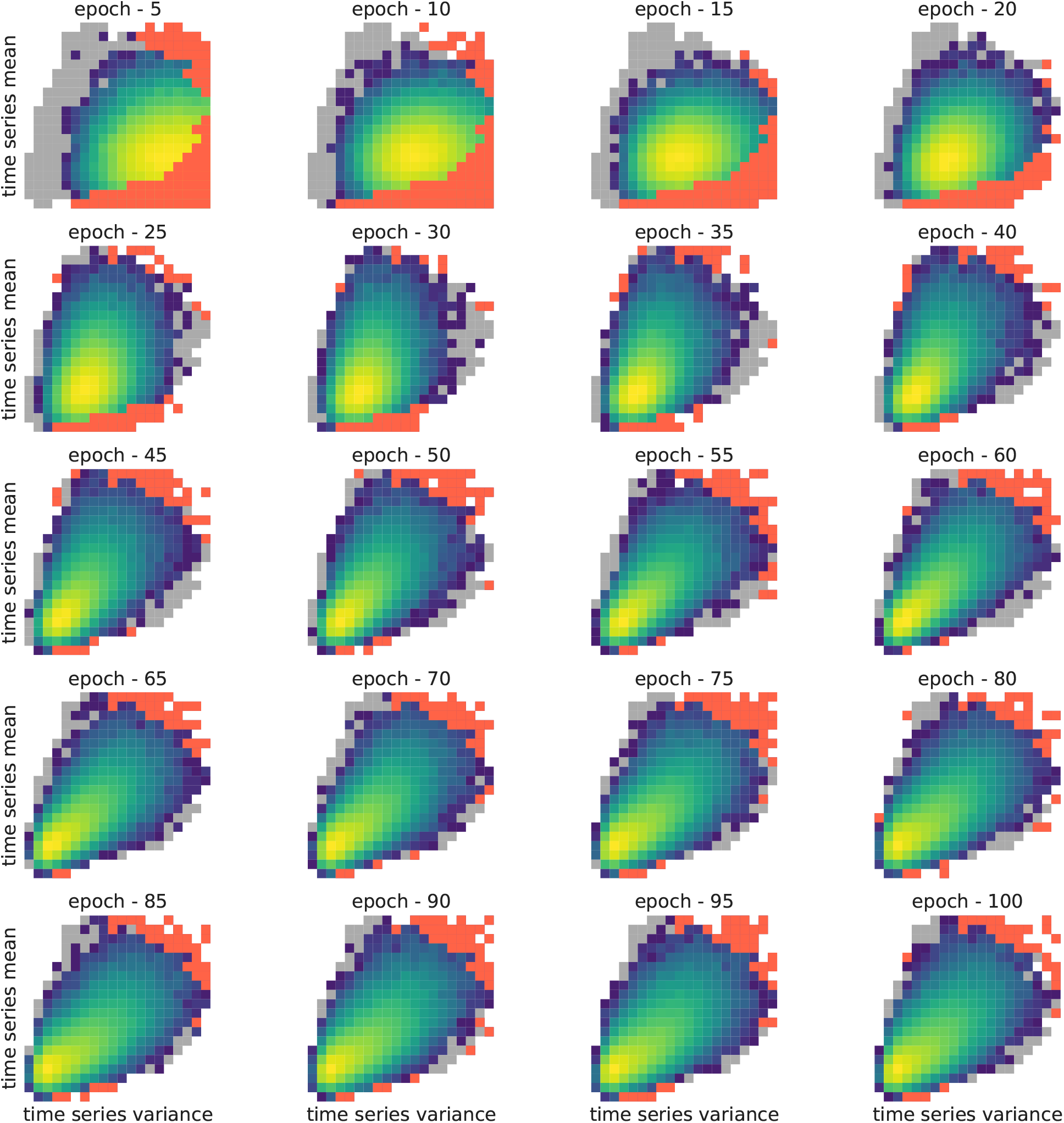
Evolution of the synthetic density of time series for the best WGAN trained on regular glucose lab time series, every 5 epochs.

**Figure 17:**
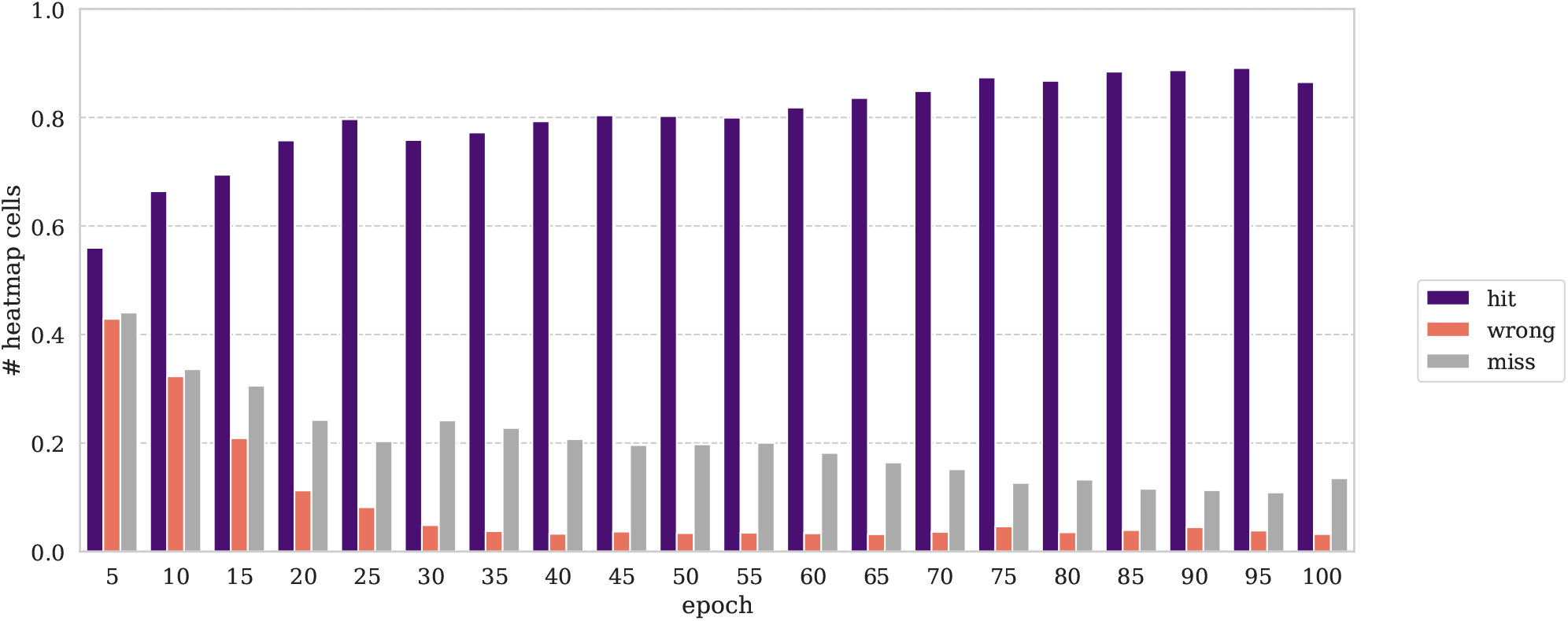
Weighted ratio of density categories covered by the best synthetic density every 5 epochs.

### 3.2 Irregular LTTS with auxiliary drug exposure generation with conditional WGAN-GP

We then proceeded to train the conditional WGAN on the irregular laboratory test time series using auxiliary drug exposures as conditional information. While generating realistic synthetic glucose time series is a good first step, the generation is only random due to the nature of GAN as implicit generative models. We do not have any control over the mode of the generated samples, the properties, or the sub-group to which these time series belong. Conditional GANs on the other hand allow for known auxiliary information to be used during the training, at the input of the generator and the critic. Once trained, the conditional GAN generator takes as an input a random vector concatenated to an auxiliary information vector. While the former is useful for diversity in the stochastic generation process, the latter enables the user to select the conditional information of the time series generated and therefore direct this generation.

#### 3.2.1 Hyperparameter tuning

Given the major similarities between the WGAN and the conditional WGAN, we only rerun the 10 best hyperparameter sets from the previous study. The model selection followed the same process, at the difference that we had to run these experiments for each drug terminology and conditional vector length (i.e., eight modalities), similarily to the forecasting experiments described in the *Supplementary Material*: ATC-3, ATC-4, ATC-5 and RxNorm with 5 or 10 drug concepts. Because RxNorm with 5 drugs yielded the most realistic conditional WGAN, the figures illustrating the following analyses are for that dataset only. The figures associated to the other drug terminologies and conditional vector lengths can be found in the Appendix (Figures **??,??,??,??,??,??,??**).

#### 3.2.2 Impact of sampling on FID

The same phenomenon than in the previous section was observed: the FID was stable through sampling rate, and only the standard deviation of the FID decreased as the sample size of the generated data increased (Figure 18). The real training set and the testing set had an FID of 11.04 (mean diff.: 1.06, cov. diff.: 9.98) while the FID with the synthetic data ranged from 15.67 to 43.26 across the various drug representation tested.

**Figure 18:**
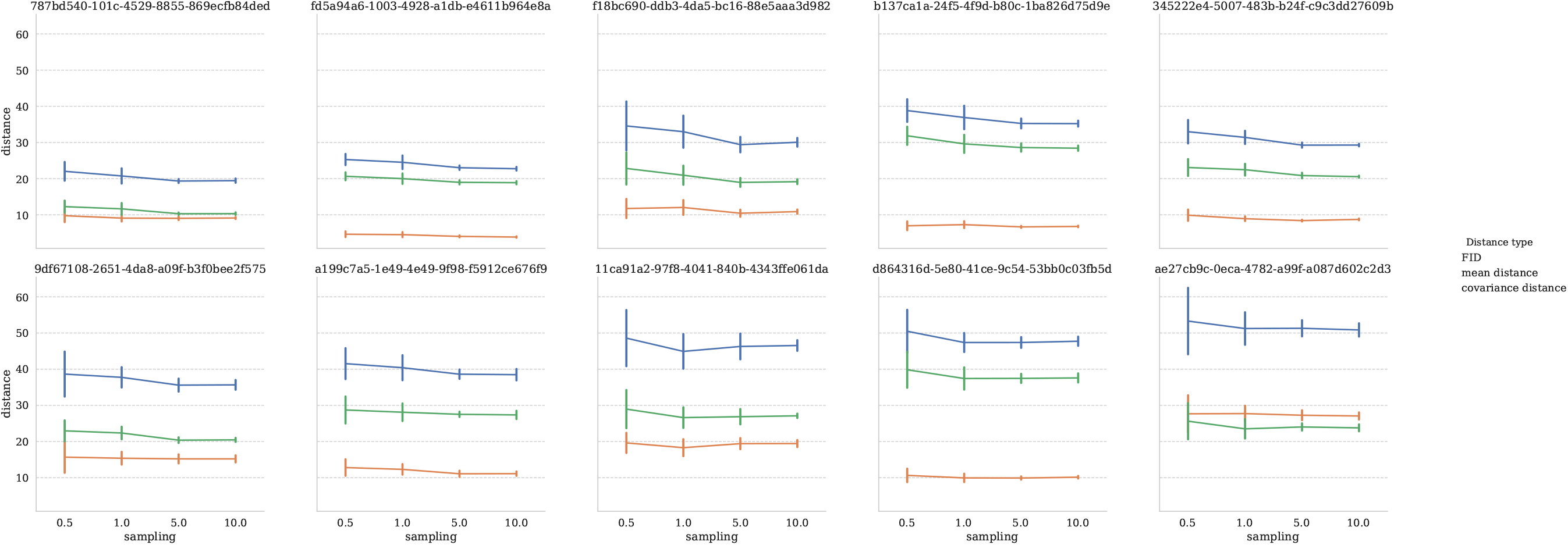
Evolution of the FID for different amounts of sampled synthetic data, expressed in ratio of real data, for conditional WGANs on irregular glucose lab time series with auxiliary RxNorm 5 drug exposures.

#### 3.2.3 FID evolution during training

Similarly to the WGAN analysis, we observe that the covariance difference is the main driver of the FID, and usually decreases almost linearly, while the mean error is much lower and therefore fluctuates more with less impact on the total distance (Figure 19).

**Figure 19:**
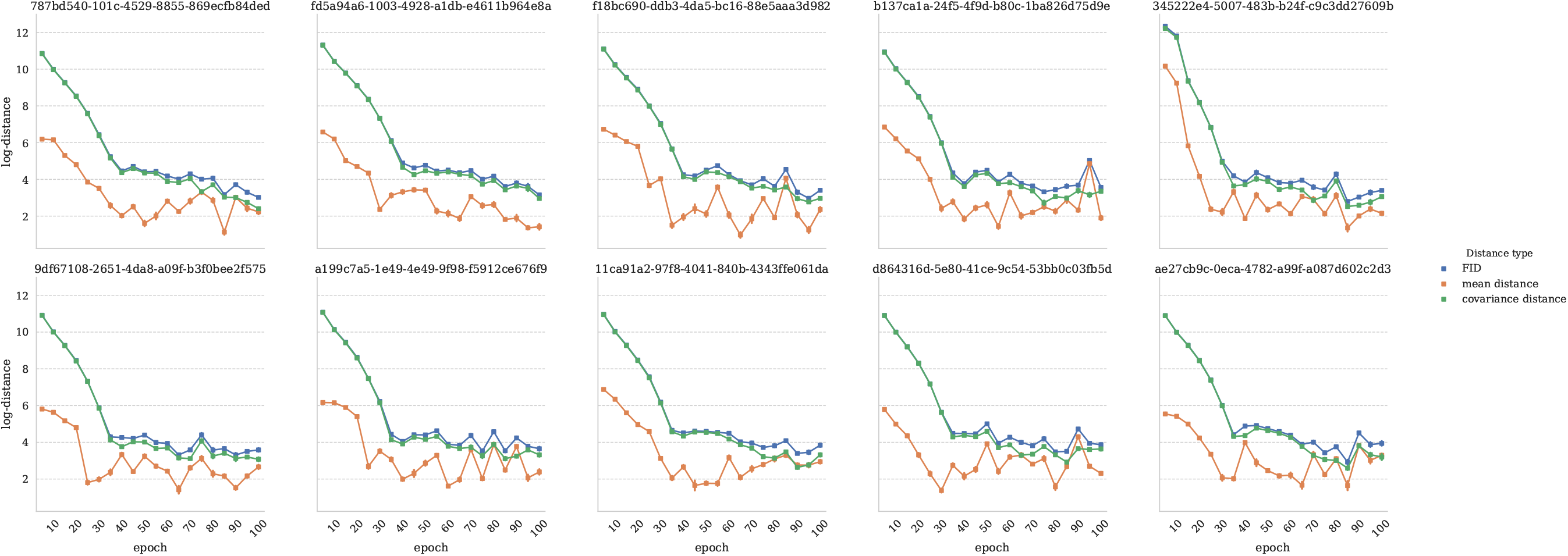
Evolution of the FID during training of WGANs on irregular glucose lab time series with RxNorm-5 auxiliary drug information, every 5 epochs.

#### 3.2.4 TSTR with a a conditional MLP forecasting model: evaluating different drug representations

Table 13 presents the best models in the TSTR task for each drug representation. The MSE on the testing set spanned from 1088.36 for ATC-5 with 5 drugs, down to 1056.32 for RxNorm with 5 drugs, effectively performing as well as a linear regression trained on the real dataset with RxNorm concepts for the top-5 drugs of exposure. ATC-5 with 5 drugs had the worst TSTR score with its best MSE at 1088.36. The FID of these conditional GANs spanned from 17.46 for RxNorm with 10 concepts, to 34.03 for ATC-3 with 10 concepts. For reference, the best non-conditional model in terms of FID scored at 26.71. The synthetic samples from conditional models were therefore overall more realistic, at the exception of ATC-3 with 10 concepts, and ATC-5 with 5 concepts. This might explain their high MSE in the TSTR task. While the best MSE with non-conditional models was 1103.84, all the TSTR MSEs of the conditional models were between 1056.32 and 1088.36.

**Table 13:** Train on Synthetic Test on Real (TSTR) evaluation of the best WGAN models by drug representation.

| Drug concepts | avg. FID (±SD) | avg. mean diff. (±SD) | avg. cov. diff (±SD) | best synth. err. | avg synth. err. (±SD) | best real training err. | avg. real train err. (±SD) | best real test err. | avg. real test err. (±SD) |
| --- | --- | --- | --- | --- | --- | --- | --- | --- | --- |
| ATC-3 5 | 18.87<br>(± 1.493) | 3.61<br>(± 1.068) | 15.27<br>(± 2.240) | 994.56 | 999.60<br>(± 5.775) | 1048.41 | 1053.19<br>(± 4.505) | 1071.34 | 1076.47<br>(± 5.740) |
| ATC-3 10 | 34.03<br>(± 2.795) | 16.71<br>(± 1.667) | 17.32<br>(± 1.376) | 933.32 | 937.66<br>(± 3.152) | 1053.22 | 1059.49<br>(± 4.224) | 1075.37 | 1082.24<br>(± 3.589) |
| ATC-4 5 | 22.62<br>(± 1.329) | 5.16<br>(± 0.706) | 17.47<br>(± 1.121) | 1038.67 | 1051.27<br>(± 14.812) | 1052.11 | 1064.65<br>(± 13.926) | 1080.41 | 1093.31<br>(± 15.487) |
| ATC-4 10 | 26.72<br>(± 1.881) | 5.90<br>(± 0.807) | 20.82<br>(± 1.345) | 982.54 | 991.07<br>(± 8.720) | 1049.31 | 1054.47<br>(± 5.185) | 1071.13 | 1079.68<br>(± 6.185) |
| ATC-5 5 | 29.14<br>(± 1.714) | 9.48<br>(± 0.960) | 19.67<br>(± 1.279) | 960.45 | 971.39<br>(± 10.017) | 1063.21 | 1079.68<br>(± 14.125) | 1088.36 | 1105.87<br>(± 13.420) |
| ATC-5 10 | 21.68<br>(± 3.147) | 5.29<br>(± 1.088) | 16.39<br>(± 2.313) | 979.75 | 984.84<br>(± 3.637) | 1041.72 | 1047.21<br>(± 4.393) | 1069.46 | 1078.61<br>(± 5.241) |
| **RxNorm 5** | 20.52<br>(± 1.283) | 9.41<br>(± 1.279) | 11.11<br>(± 0.817) | **924.91** | 929.36<br>(± 2.970) | **1035.14** | 1038.85<br>(± 3.154) | **1056.32** | 1061.85<br>(± 4.321) |
| RxNorm 10 | 17.46<br>(± 2.405) | 5.07<br>(± 1.051) | 12.39<br>(± 1.853) | 988.87 | 992.51<br>(± 3.864) | 1047.23 | 1055.96<br>(± 9.513) | 1065.80 | 1077.73<br>(± 8.408) |

#### 3.2.5 Density evolution through training of the best conditional WGAN

The best conditional GAN with 5 RxNorm concepts as auxiliary information presented an analogous behavior during training as observed in the non-conditional GAN: The learned distribution first adjusted its mean, and then its variance and covariance (Figure 20). The analysis of the coverage, wrong modes, and missed modes (Figure 21) showed a peak coverage of 92.07% at epoch 100.

**Figure 20:**
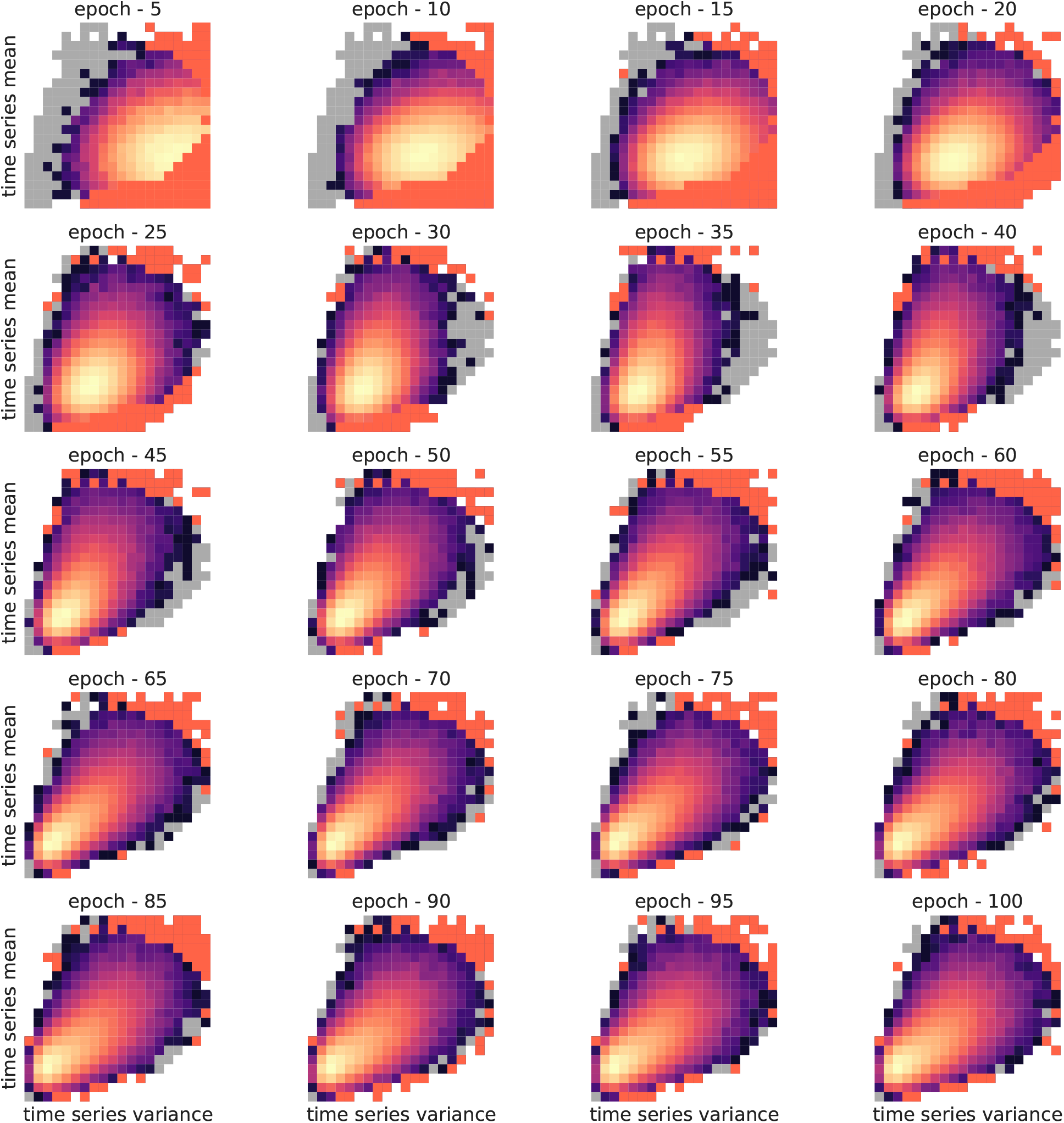
Evolution of the synthetic density of time series for the best conditional WGAN trained on irregular glucose lab time series with RxNorm-5 auxiliary drug information, every 5 epochs.

**Figure 21:**
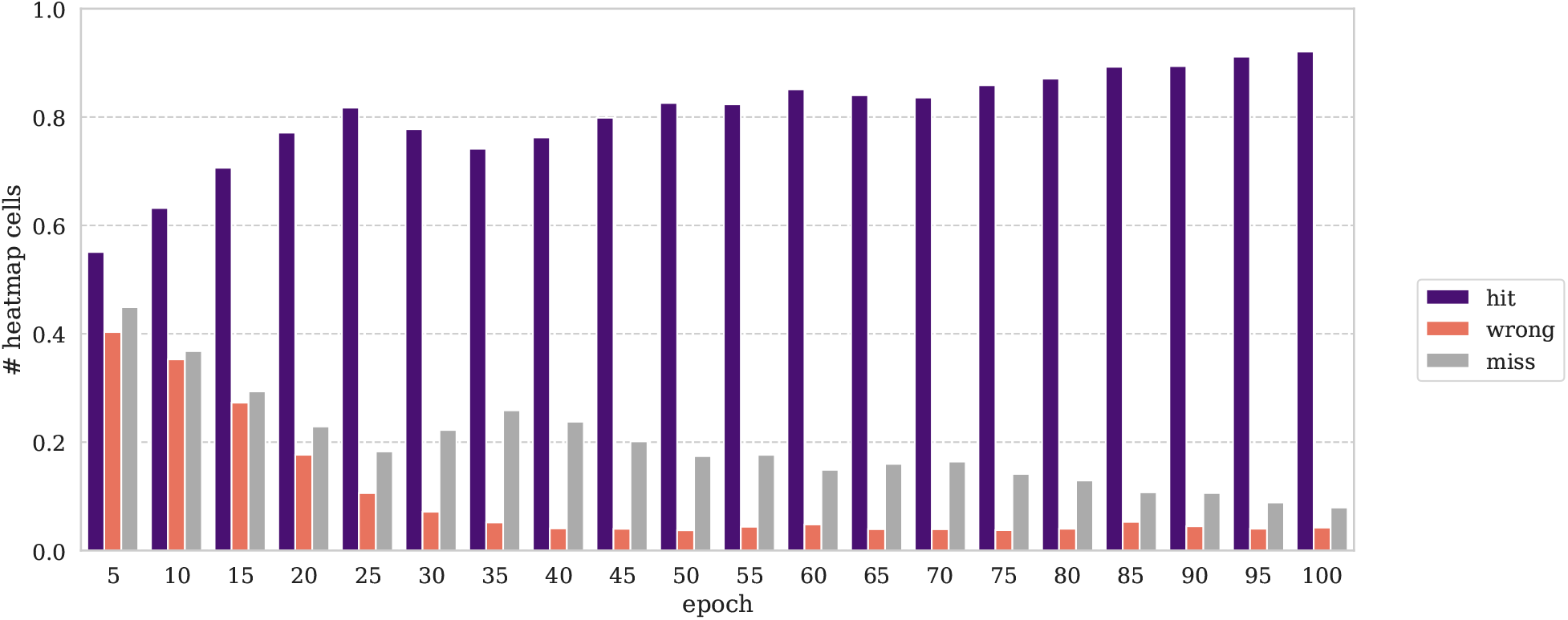
Weighted ratio of density categories covered by the best conditional WGAN trained on irregular glucose lab time series with RxNorm-5 auxiliary drug information, every 5 epochs.

### 3.3 Inferring glucose time series with drug exposures unseen at training

The original dataset contained 11,746 glucose time series exposed to a single drug at a time. These were removed from the training set that was left with 64,857 samples. the conditional WGAN was trained with the hyperparameters that yielded the best FIDs in Aim 2, and the best model out of 10 runs was selected for the simulation experiment (FID: 22.31). Let’s call this conditional WGAN the *inference* model.

For each of the 10 drugs, we generated 10,000 synthetic samples with a conditional vector incrementally increasing between 0.0 and 1.0 by steps of 0.1 for that drug of interested, and computed at every step the FID between these synthetic samples and the real samples only exposed to that drug. In order to get FID ranges of reference, we also computed the FID between synthetic samples and real single-drug samples using the conditional WGAN from Aim 2, that was exposed at training to samples with single drug of exposure (i.e., the *exposed* model). The FID between synthetic and real samples at different interpolation values in the latent spaces are displayed in figures 22 to 31.

**Figure 22:**
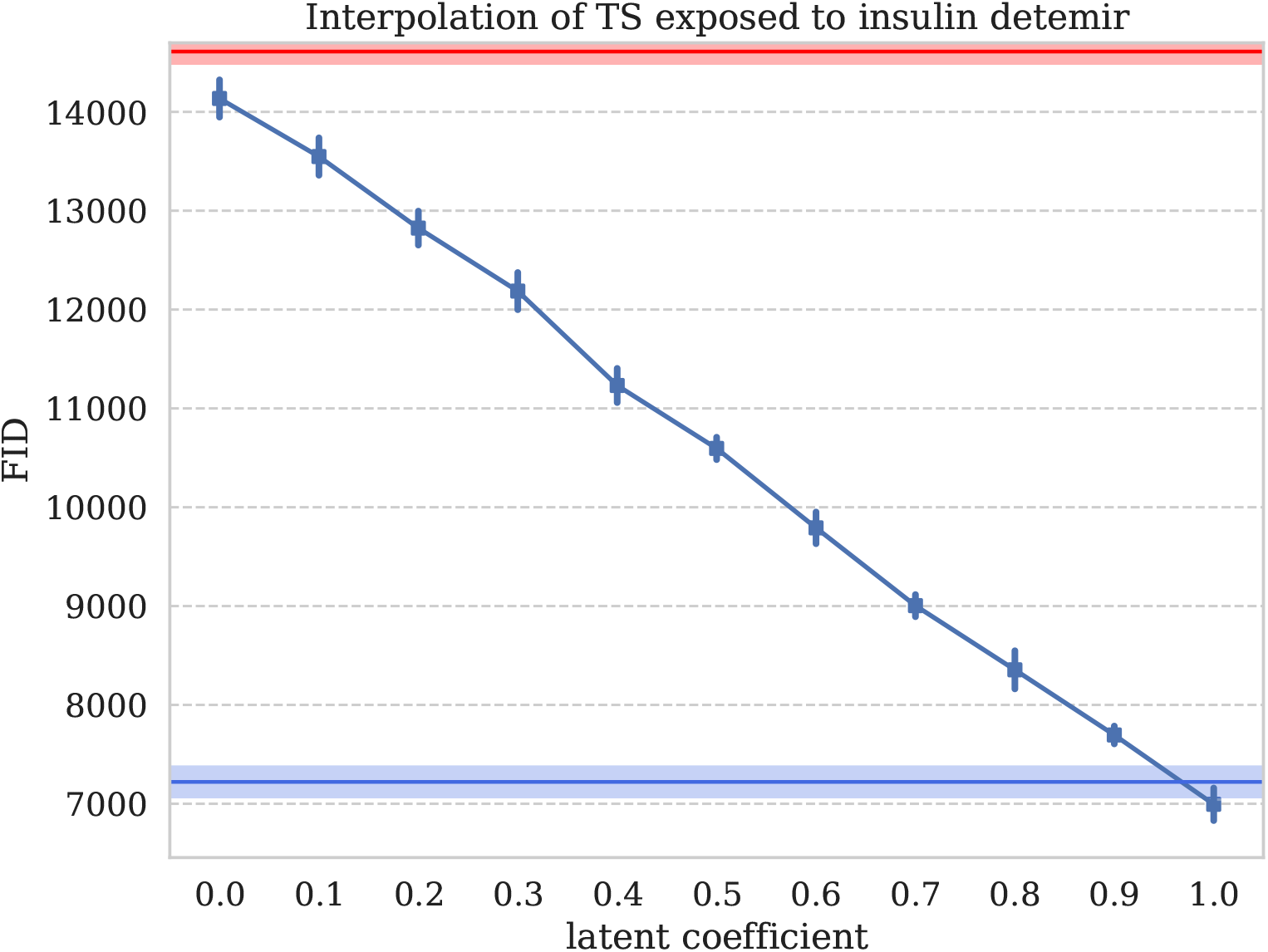
FID between synthetic and real glucose time series, for different values in the conditional vector between 0, non-exposed, and 1, exposed to insulin detemir

We observed two phenomenons: (1) sample sets generated along that interpolated conditional exposure vector presented a continuously decreasing FID with the real data, showing the interpolation power of the latent space in generative adversarial networks; (2) for all drugs, the model never exposed to single drug exposures got an FID close to the model that was trained with single-drug exposure samples. Insulin detemir exposed time series were inferred by the *inference* model with an FID comparable to the model that was exposed to it. There was one outlying drug: isopropanol, where our *inference* model samples were more realistic than the sample of the *exposed* GAN that was trained on these single exposure time series. Table 14 summarizes the FID between synthetic and real samples for the model exposed at training, and the model not exposed at training.

**Table 14:** FID between real and synthetic data for exposure to a unique drug at a time: *inference* conditional WGAN never exposed to these single-exposure samples, and *exposed* conditional WGAN that was trained with samples exposed to these unique drug exposures.

| drug of exposure | inference WGAN FID (± SD) | exposed WGAN FID (± SD) |
| --- | --- | --- |
| insulin detemir | 6994.51 (± 173.328) | 7221.17 (± 189.009) |
| sitagliptin | 1770.57 (± 49.856) | 589.87 (± 25.828) |
| insulin glargine | 952.76 (± 76.363) | 249.47 (± 11.458) |
| insulin lispro | 5700.67 (± 84.937) | 2094.06 (± 91.808) |
| metformin | 643.97 (± 37.603) | 249.45 (± 19.172) |
| isopropanol | 1184.67 (± 61.595) | 2051.22 (± 63.224) |
| glipizide | 1127.71 (± 41.407) | 478.10 (± 11.196) |
| glimepiride | 2049.26 (± 55.207) | 1757.40 (± 71.324) |
| glibenclamide | 2099.16 (± 84.242) | 1094.61 (± 41.500) |
| insulin (human) | 359.62 (± 29.739) | 66.40 (± 8.364) |

For visualization purposes, we represented these single-exposure samples for the inference model, the expose model, and the real data density represented by variance and mean of the time series (Figures 32, 33).

**Figure 23:**
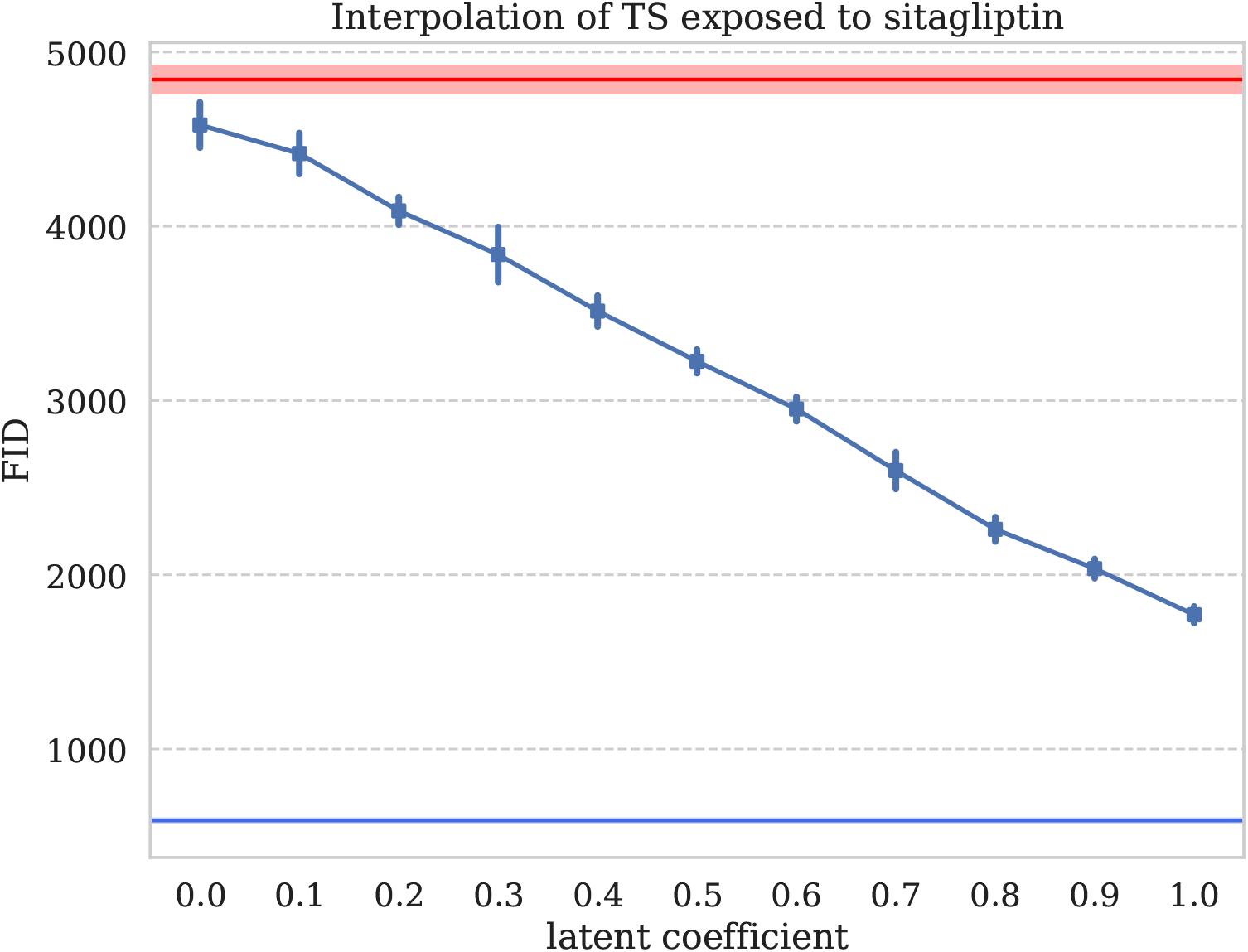
FID between synthetic and real glucose time series, for different values in the conditional vector between 0, non-exposed, and 1, exposed to sitagliptin

**Figure 24:**
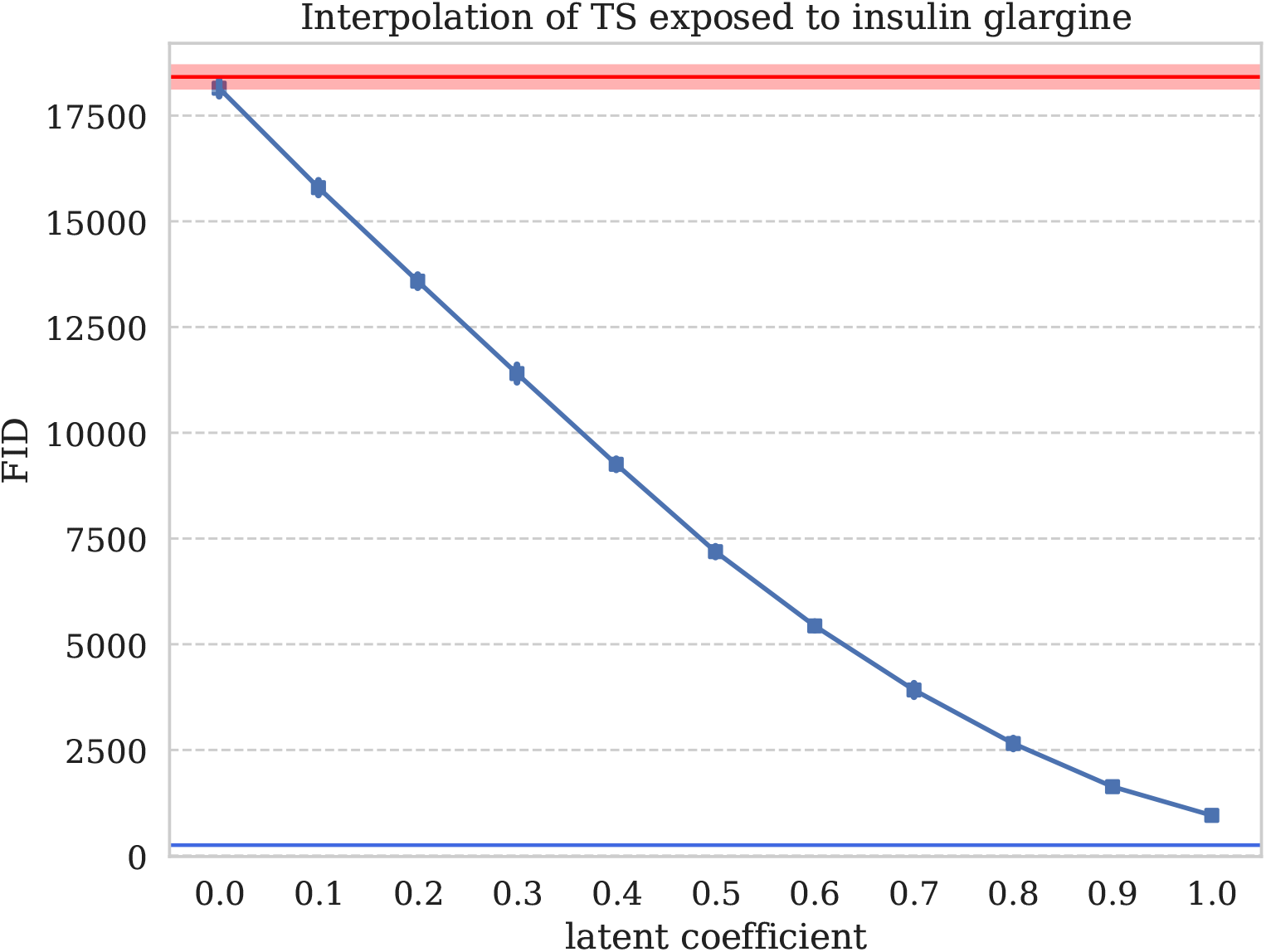
FID between synthetic and real glucose time series, for different values in the conditional vector between 0, non-exposed, and 1, exposed to insulin glargine

**Figure 25:**
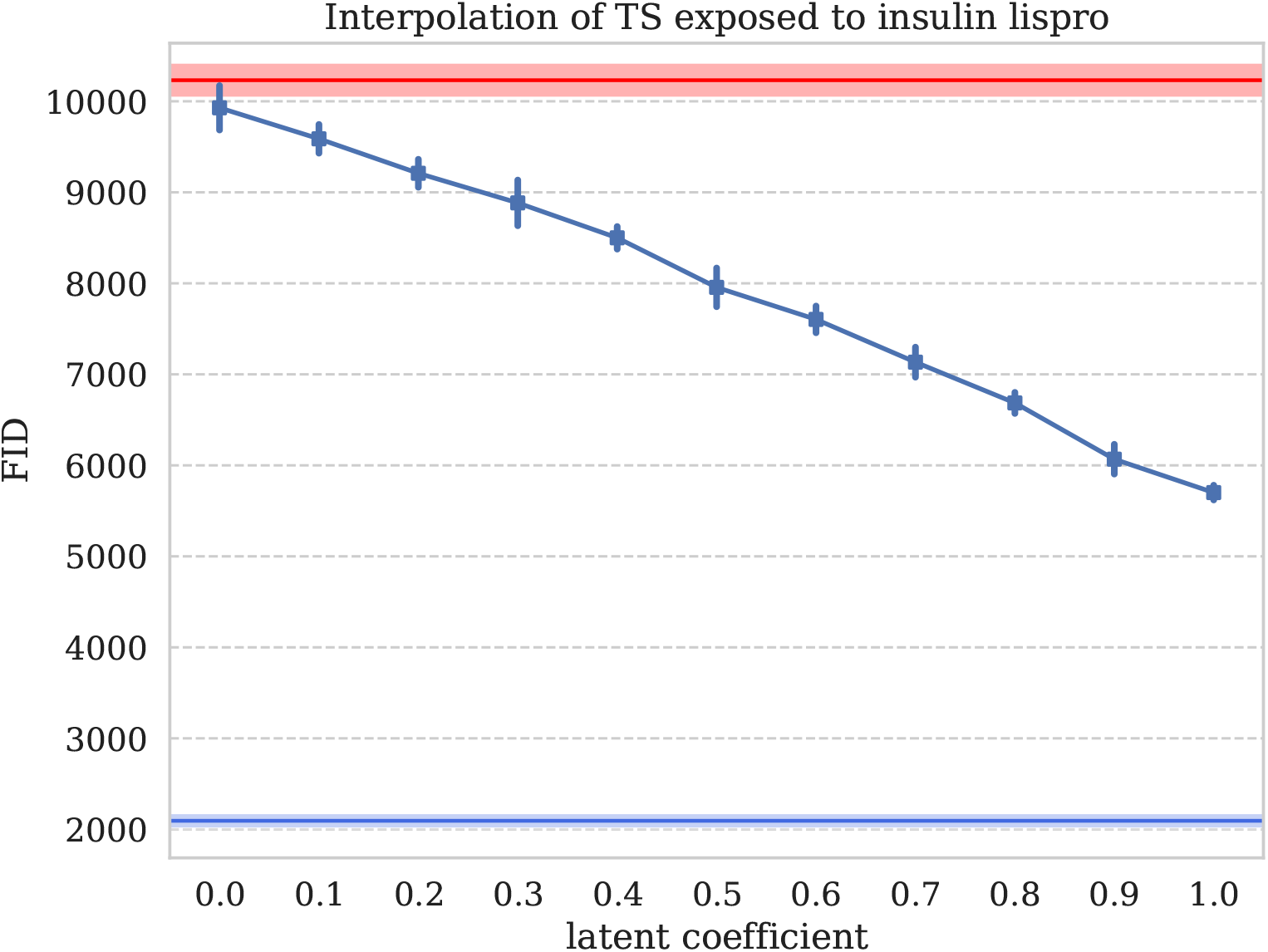
FID between synthetic and real glucose time series, for different values in the conditional vector between 0, non-exposed, and 1, exposed to insulin lispro

**Figure 26:**
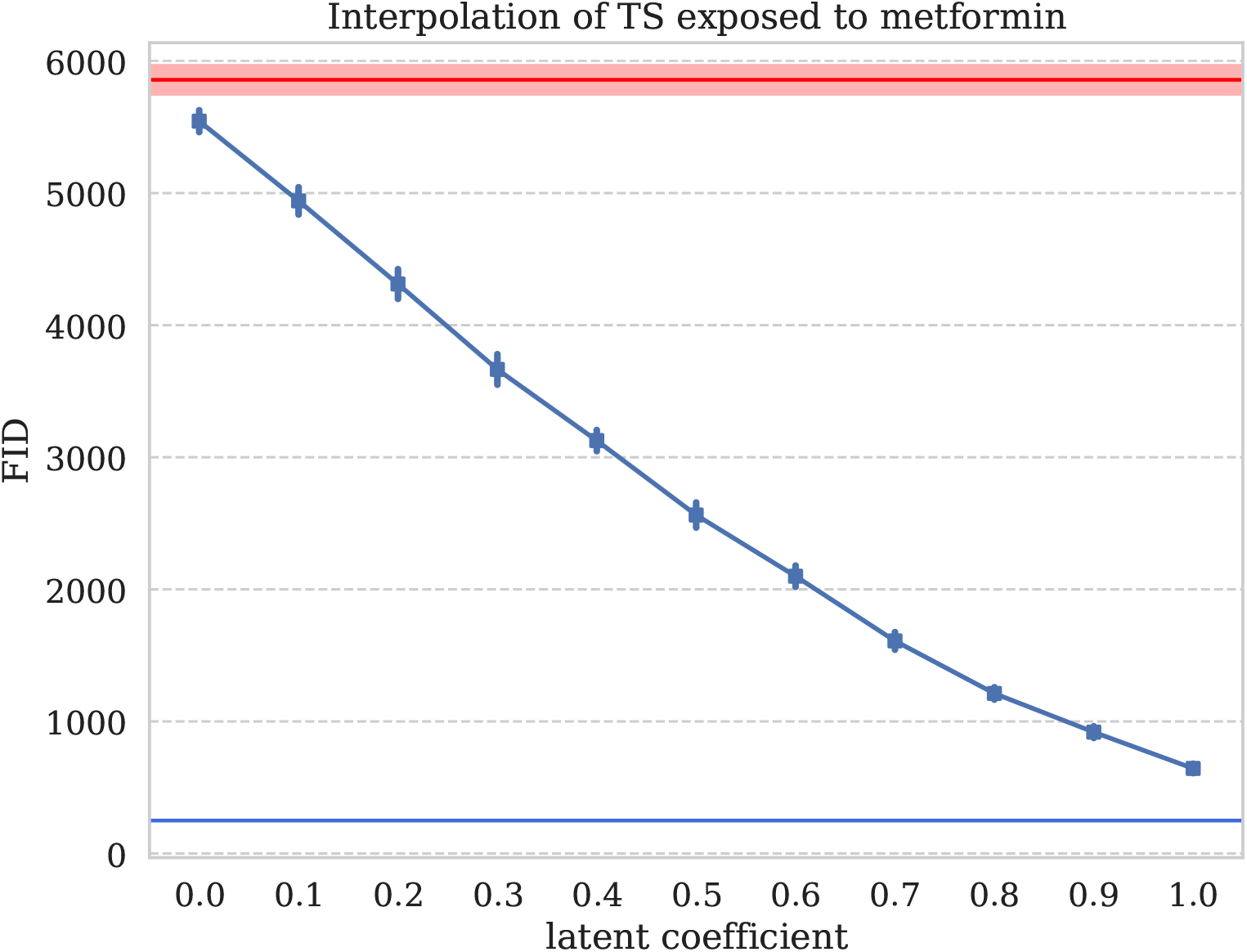
FID between synthetic and real glucose time series, for different values in the conditional vector between 0, non-exposed, and 1, exposed to metformin

**Figure 27:**
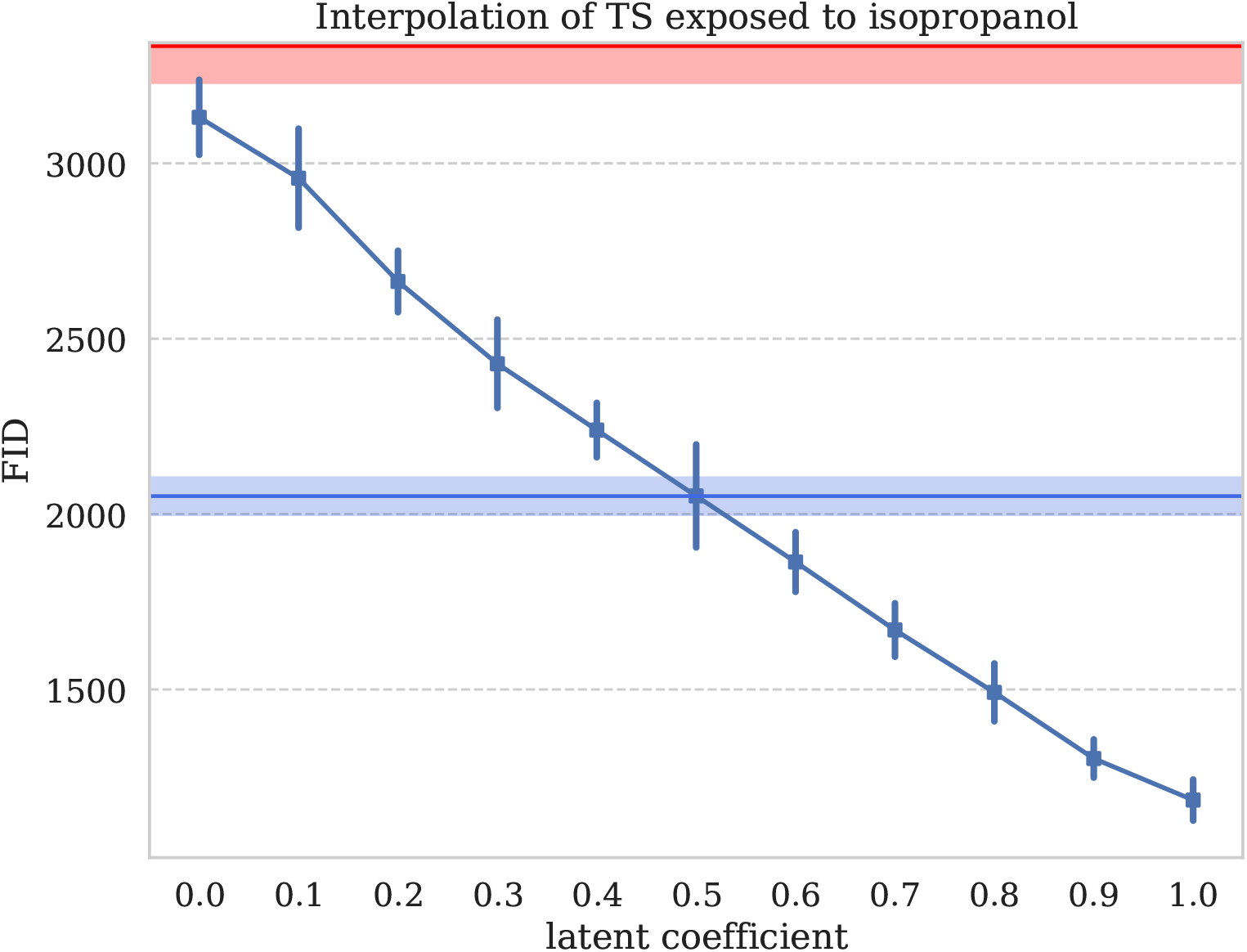
FID between synthetic and real glucose time series, for different values in the conditional vector between 0, non-exposed, and 1, exposed to isopropanol

**Figure 28:**
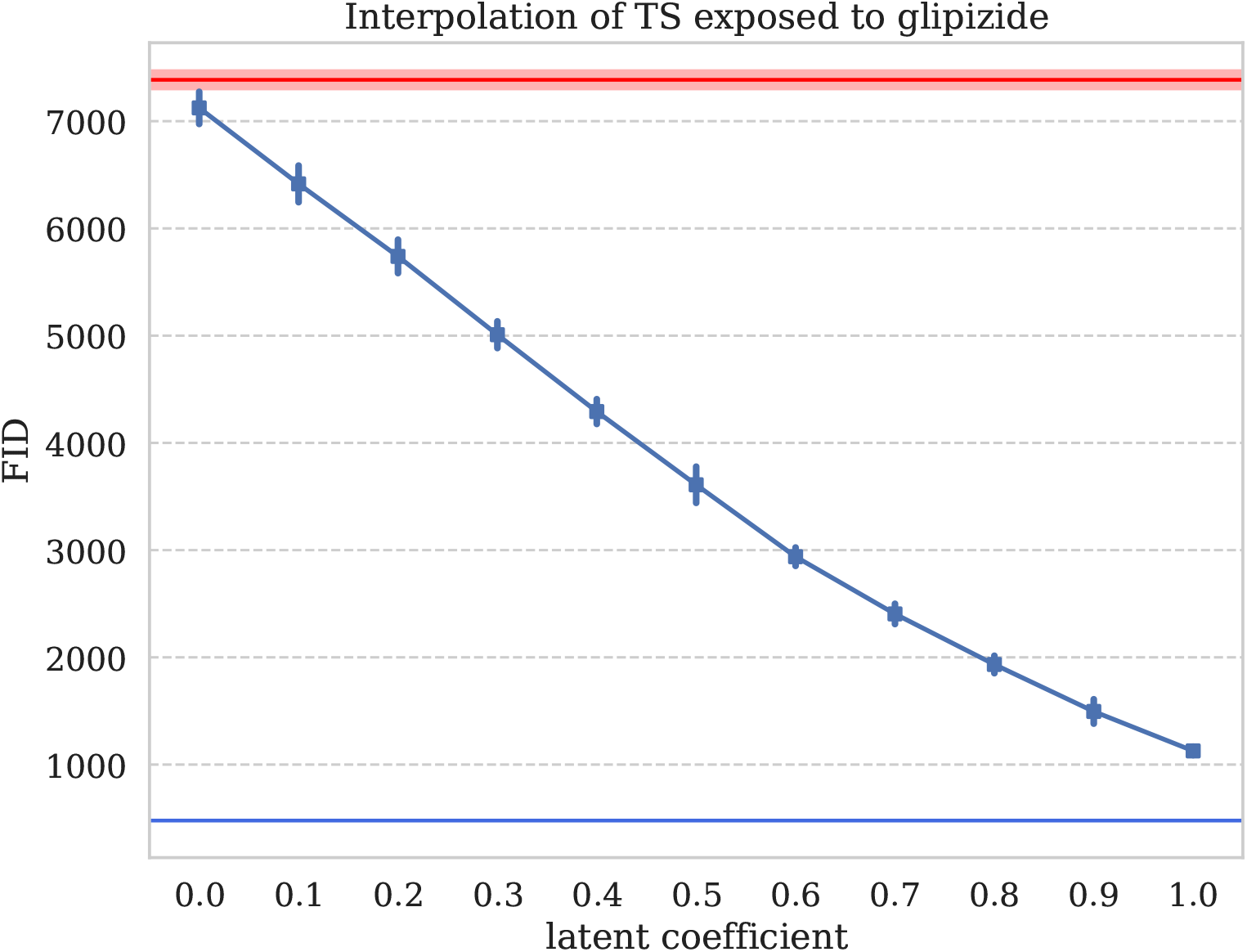
FID between synthetic and real glucose time series, for different values in the conditional vector between 0, non-exposed, and 1, exposed to glipizide

**Figure 29:**
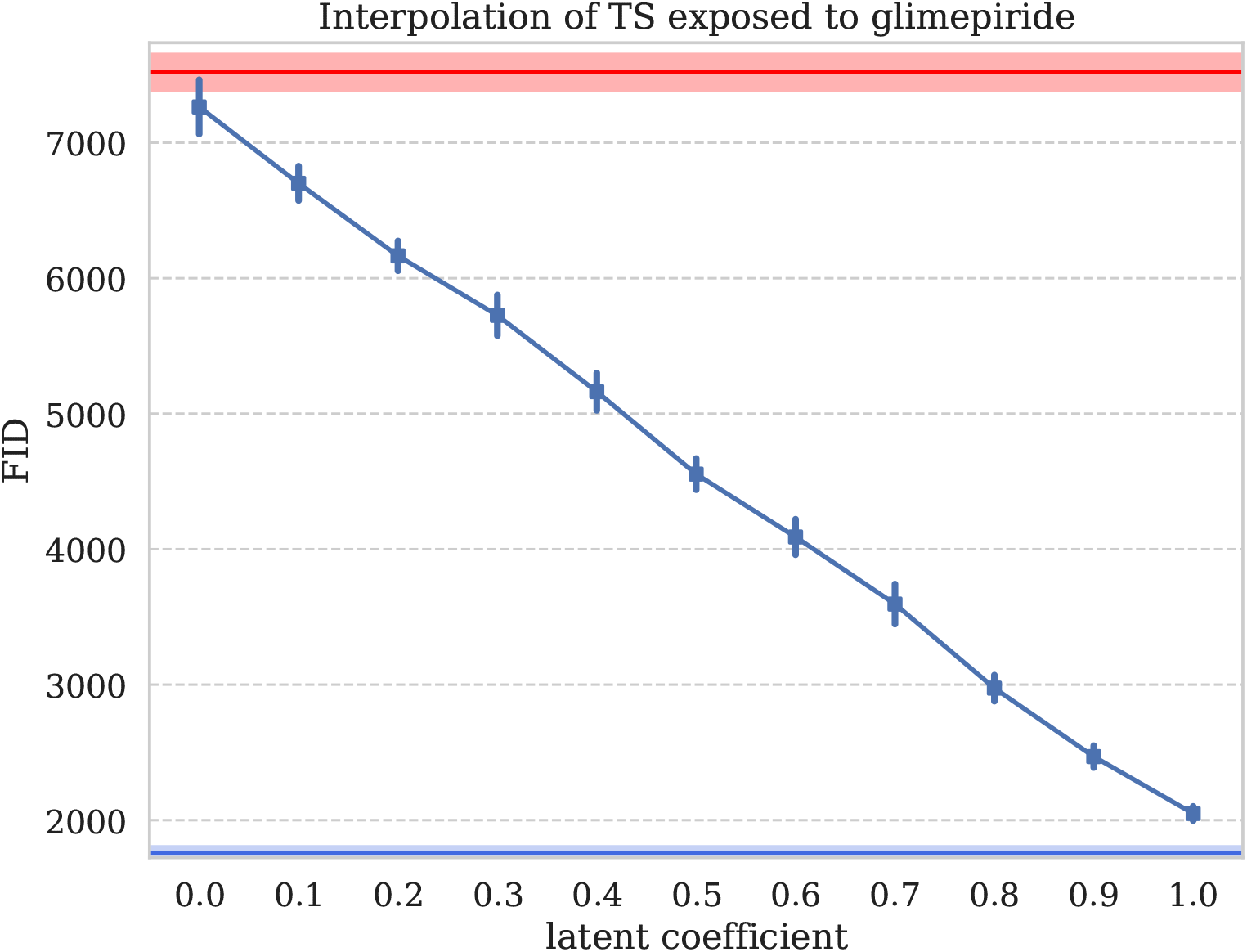
FID between synthetic and real glucose time series, for different values in the conditional vector between 0, non-exposed, and 1, exposed to glimepiride

**Figure 30:**
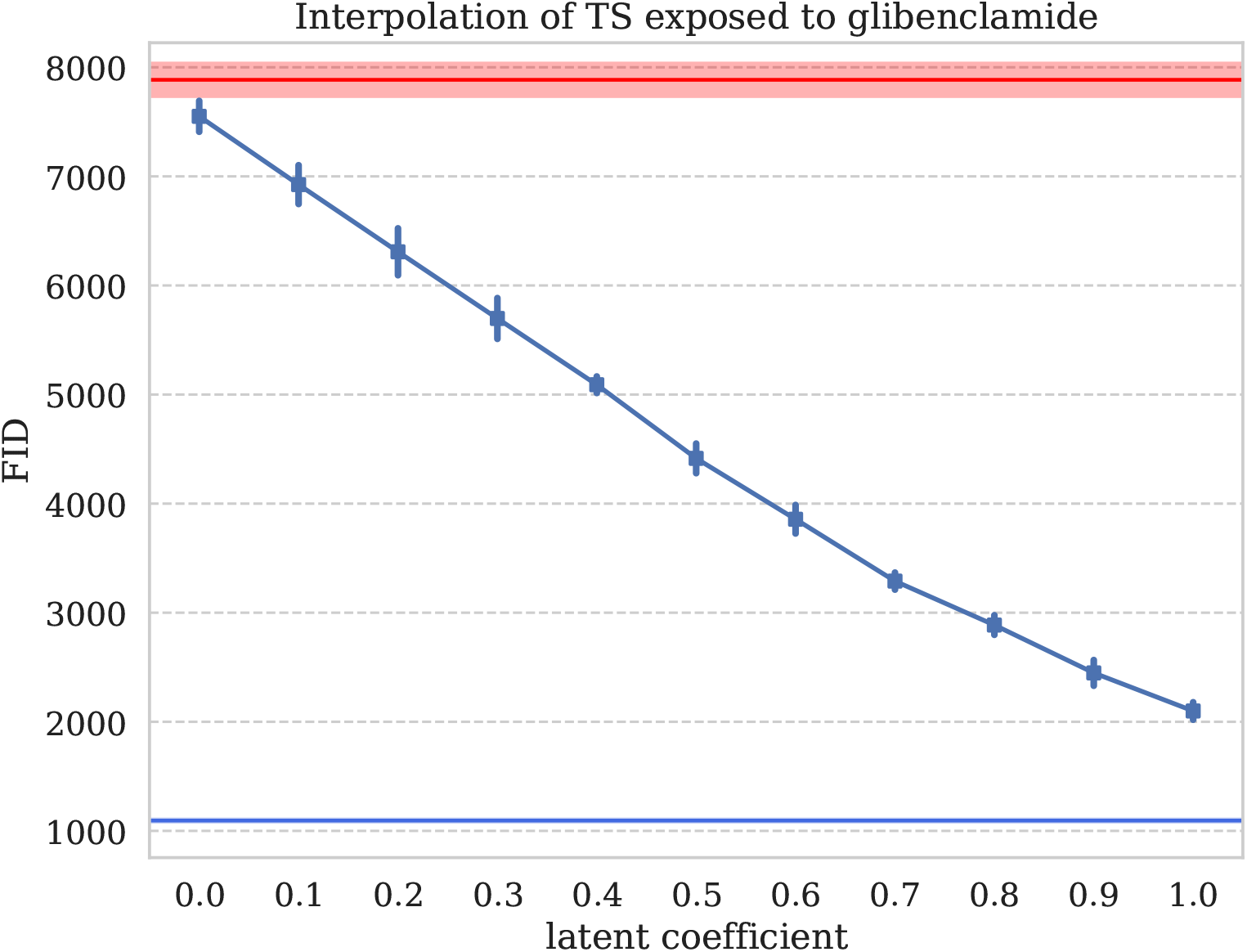
FID between synthetic and real glucose time series, for different values in the conditional vector between 0, non-exposed, and 1, exposed to glibenclamide

**Figure 31:**
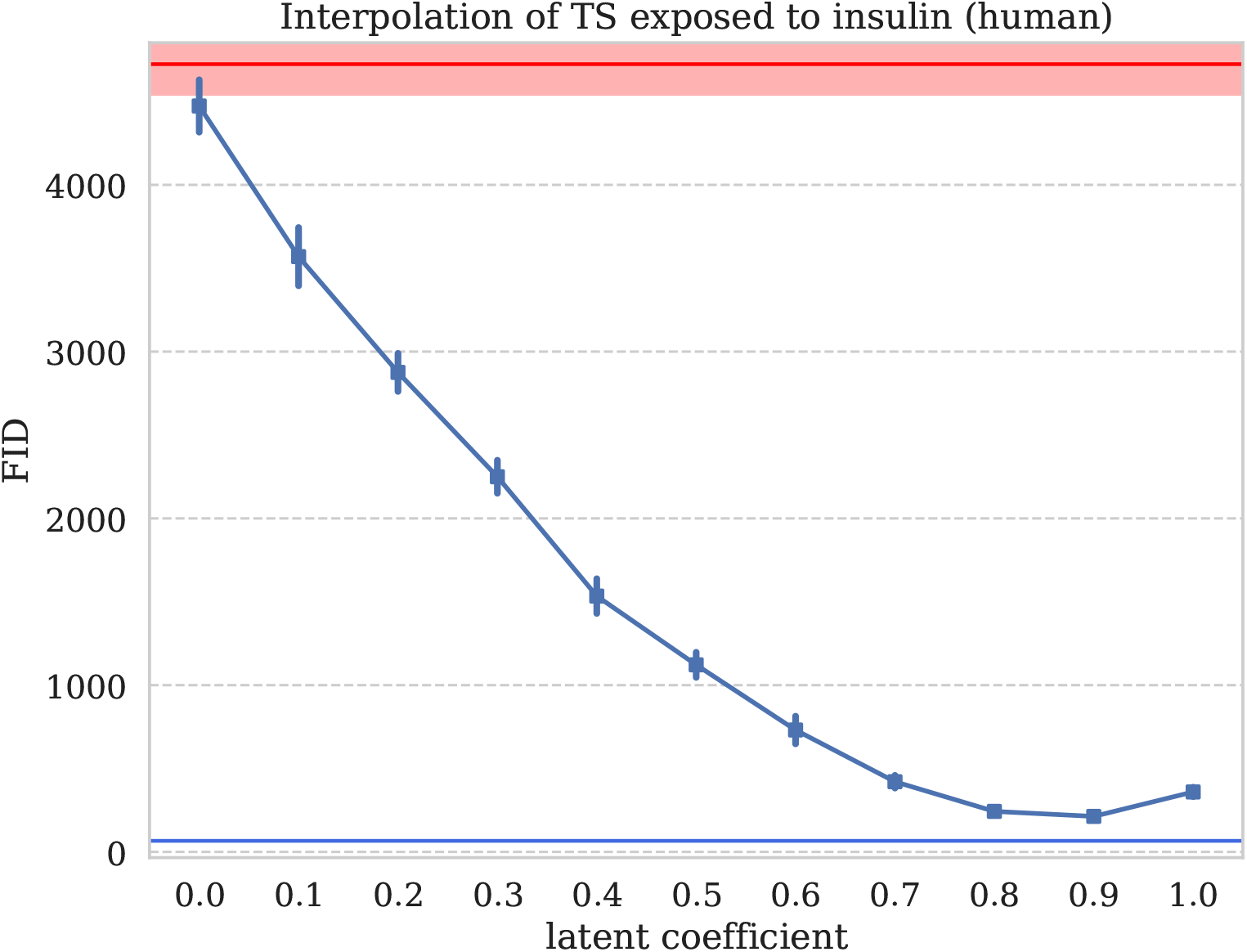
FID between synthetic and real glucose time series, for different values in the conditional vector between 0, non-exposed, and 1, exposed to insluin (human)

**Figure 32:**
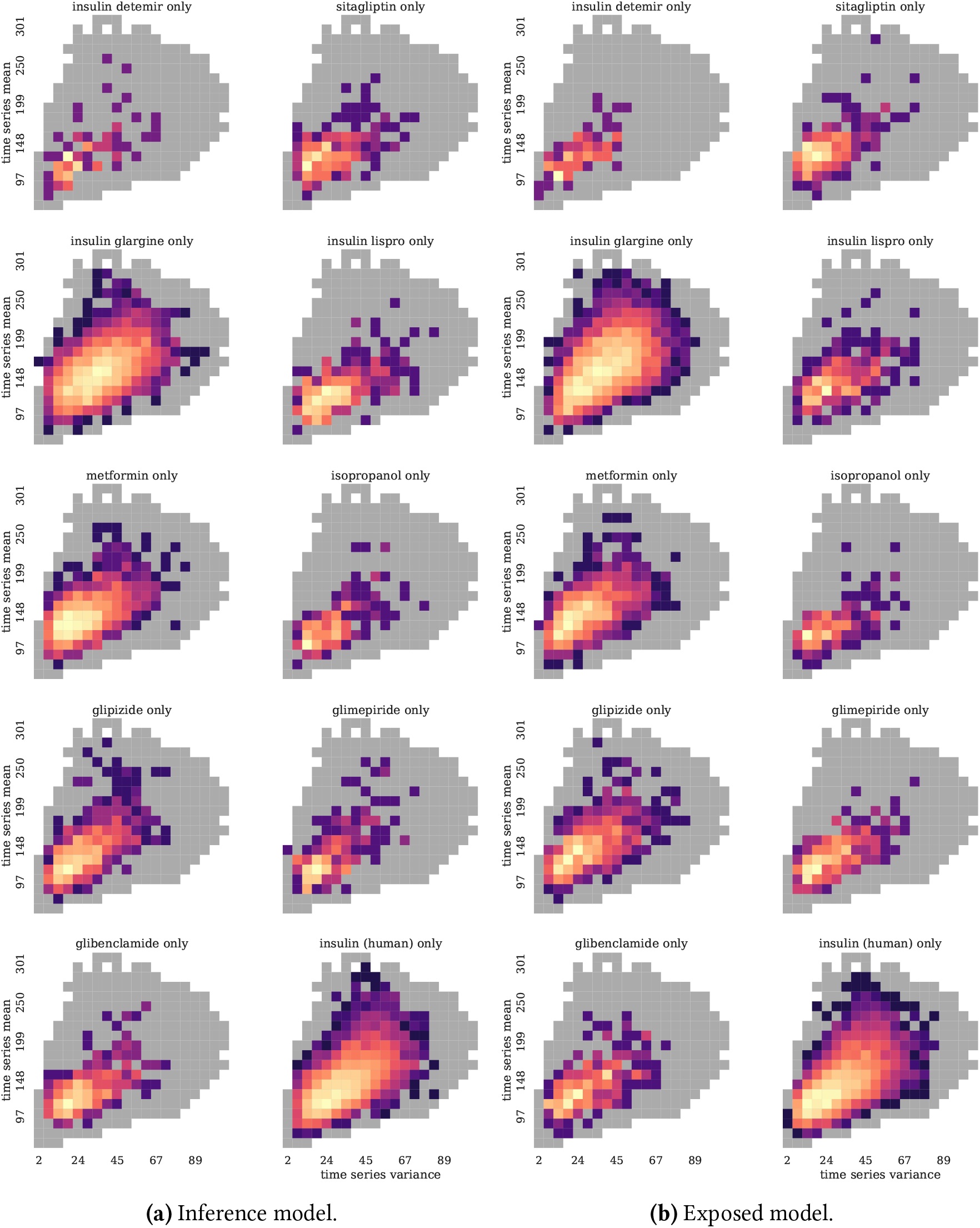
Density heatmap of irregular glucose lab time series exposed to the top-10 RxNorm, synthetic vs. real with equal sampling.

**Figure 33:**
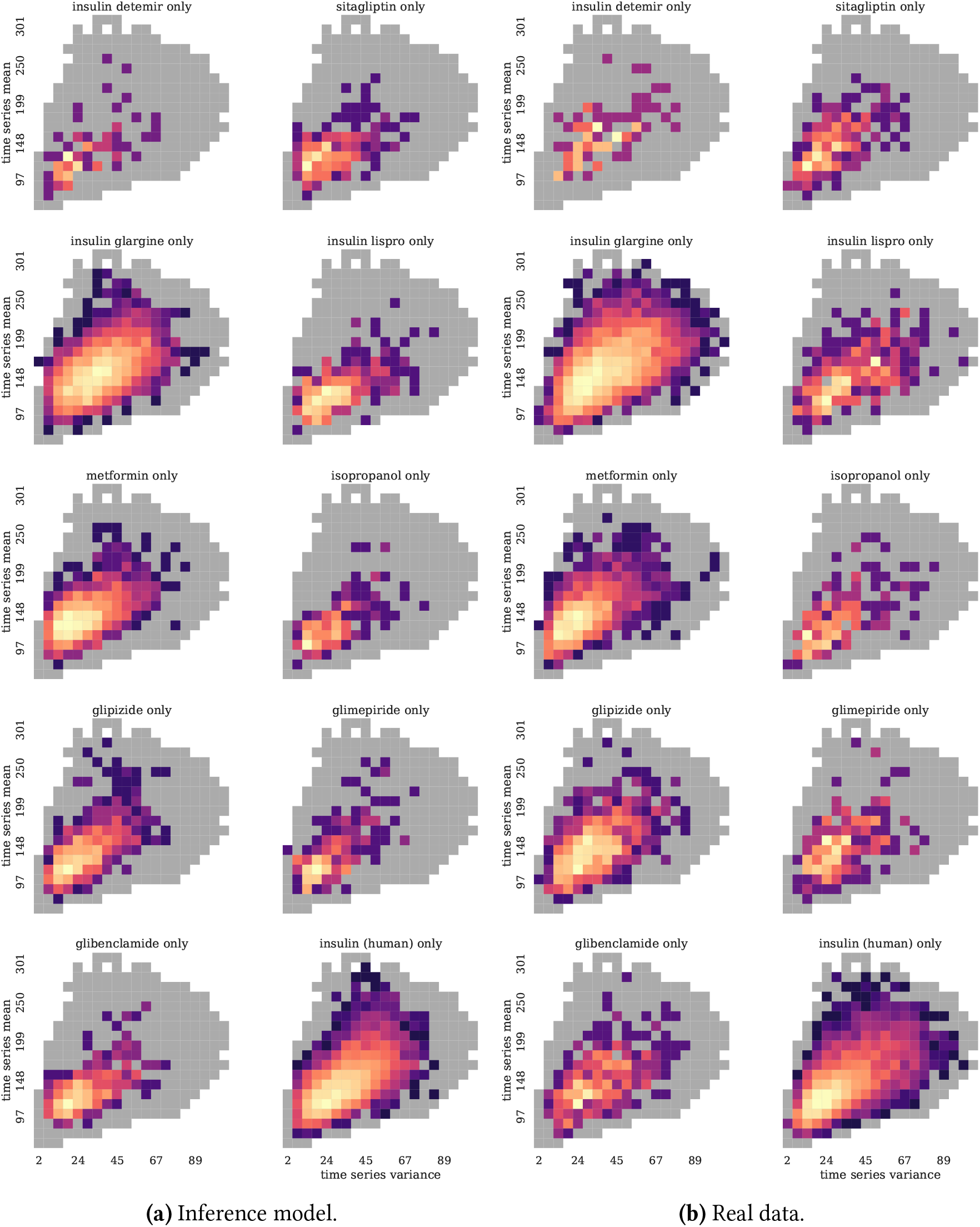
Density heatmap of irregular glucose lab time series exposed to the top-10 RxNorm, synthetic vs. real with fixed synthetic sampling at 10% of the training set size.

### 3.4 Targeted data augmentation with conditional WGANs

Now that we demonstrated that conditional WGAN can generate time series with conditional information that was not seen at training, we built some confidence about the ability of these models to augment real datasets. Indeed, the ideal target for data augmentation are sub-groups of samples that have low sample counts in the training set and high errors when the model is applied on the testing set. We showed that conditional WGAN can simulate glucose time series even when their belong to subgroups not seen at training, which should extend to time series with very few samples at training.

As a first experiment, we augmented the single drug exposure time series. Qyality control of the input time series seemed to be very important in the data augmentation part when adding synthetic data to real data. By applying the same criteria applied to the real data to the synthetic data, forecasting performances dramatically improved between no quality control 15, and identical quality control as in the forecasting experiments of Aim 1 with real data 16. Only drugs 1 (insulin detemir), 4 (insulin lispro) and 10 (insulin (human)) showed improvements when the training set was augmented with simulated time series exposed to these drugs only.

We grouped irregular glucose time series by drug exposures (RxNorm, 10 concepts), and represented these groups by their count in the training set and the MSE on the testing set based on the MLP forecasting model from Aim 1 (Figure 34).

**Figure 34:**
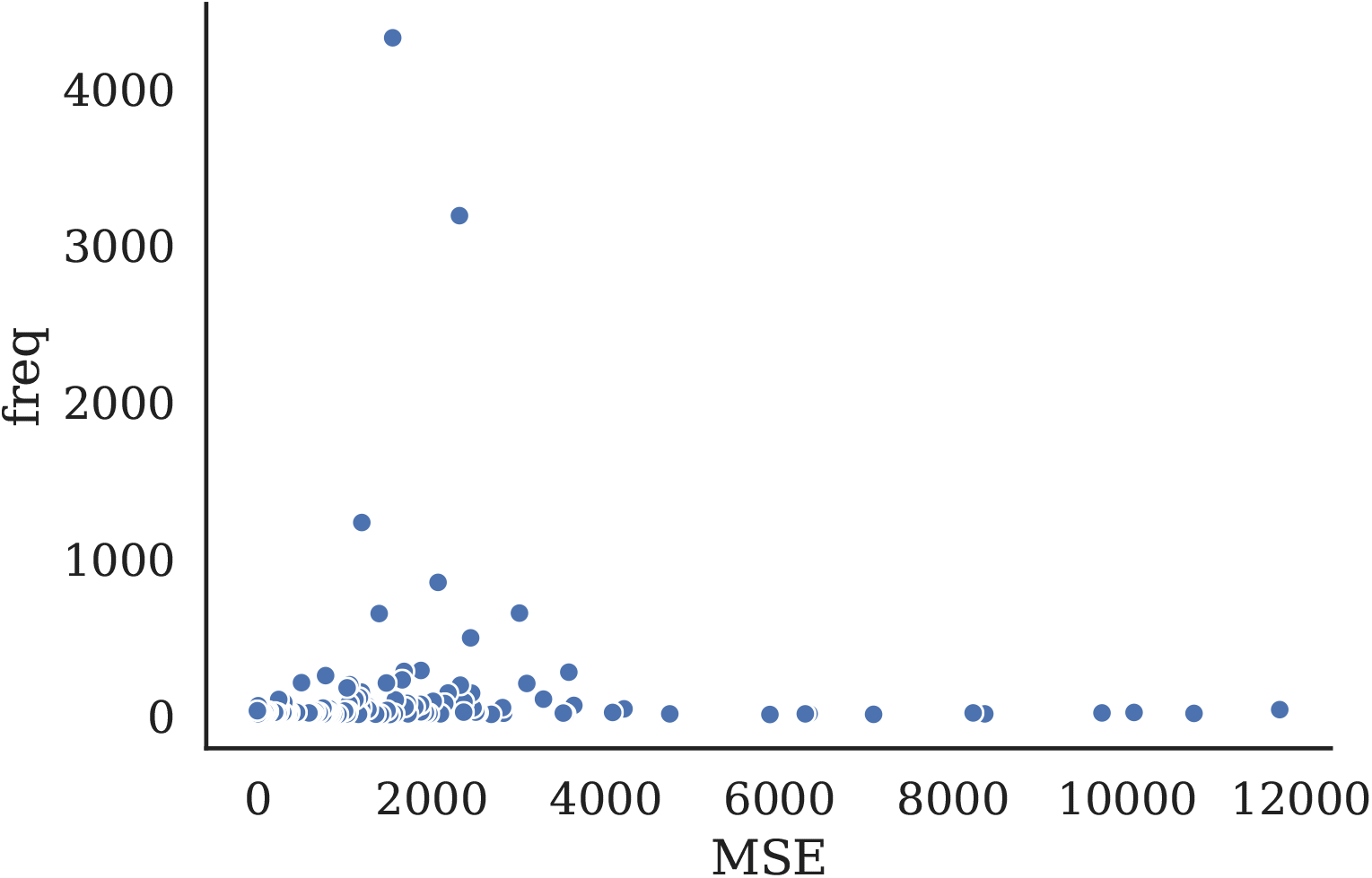
Drug exposures represented by their count in gluocose time series in the training set, and their MSE in the testing set using the best performing MLP forecasting model.

In spite of the low number of samples for these combinations, augmenting these types of glucose time series in the training set led to improved MSE in 7 out of 10 cases (Table 17).

**Table 15:**
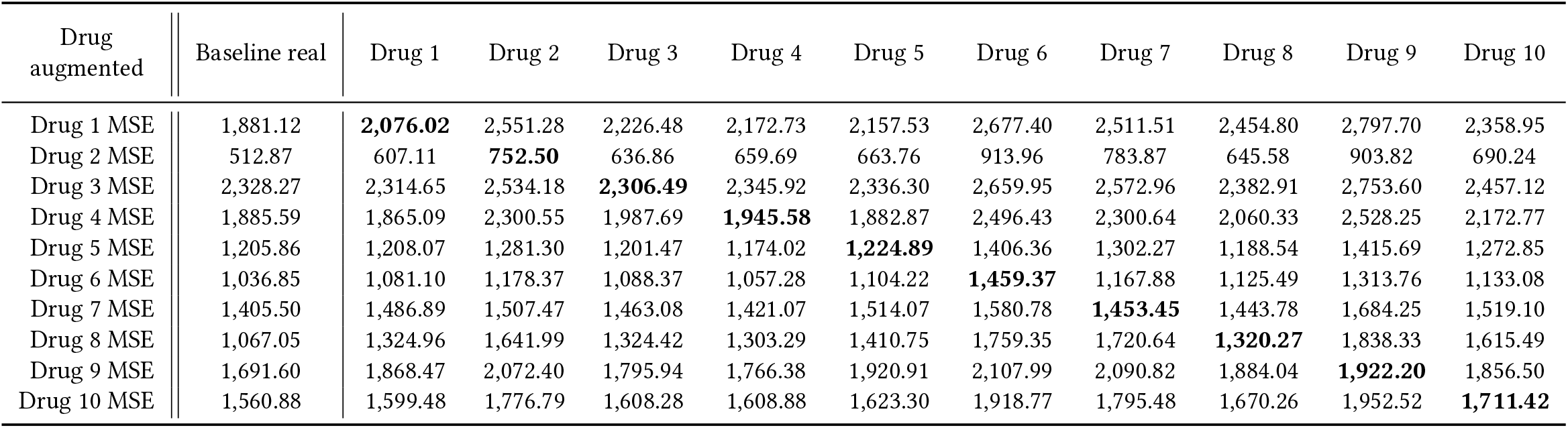
MSE by sub-group exposed to only one of the top-10 RxNorm concepts to evaluated the effects of drug-specific data augmentation of 10% of the total dataset size with a conditional WGAN. No quality control on the synthetic time series.

| Drug augmented | Baseline real | Drug 1 | Drug 2 | Drug 3 | Drug 4 | Drug 5 | Drug 6 | Drug 7 | Drug 8 | Drug 9 | Drug 10 |
| --- | --- | --- | --- | --- | --- | --- | --- | --- | --- | --- | --- |
| Drug 1 MSE | 1,881.12 | **2,076.02** | 2,551.28 | 2,226.48 | 2,172.73 | 2,157.53 | 2,677.40 | 2,511.51 | 2,454.80 | 2,797.70 | 2,358.95 |
| Drug 2 MSE | 512.87 | 607.11 | **752.50** | 636.86 | 659.69 | 663.76 | 913.96 | 783.87 | 645.58 | 903.82 | 690.24 |
| Drug 3 MSE | 2,328.27 | 2,314.65 | 2,534.18 | **2,306.49** | 2,345.92 | 2,336.30 | 2,659.95 | 2,572.96 | 2,382.91 | 2,753.60 | 2,457.12 |
| Drug 4 MSE | 1,885.59 | 1,865.09 | 2,300.55 | 1,987.69 | **1,945.58** | 1,882.87 | 2,496.43 | 2,300.64 | 2,060.33 | 2,528.25 | 2,172.77 |
| Drug 5 MSE | 1,205.86 | 1,208.07 | 1,281.30 | 1,201.47 | 1,174.02 | **1,224.89** | 1,406.36 | 1,302.27 | 1,188.54 | 1,415.69 | 1,272.85 |
| Drug 6 MSE | 1,036.85 | 1,081.10 | 1,178.37 | 1,088.37 | 1,057.28 | 1,104.22 | **1,459.37** | 1,167.88 | 1,125.49 | 1,313.76 | 1,133.08 |
| Drug 7 MSE | 1,405.50 | 1,486.89 | 1,507.47 | 1,463.08 | 1,421.07 | 1,514.07 | 1,580.78 | **1,453.45** | 1,443.78 | 1,684.25 | 1,519.10 |
| Drug 8 MSE | 1,067.05 | 1,324.96 | 1,641.99 | 1,324.42 | 1,303.29 | 1,410.75 | 1,759.35 | 1,720.64 | **1,320.27** | 1,838.33 | 1,615.49 |
| Drug 9 MSE | 1,691.60 | 1,868.47 | 2,072.40 | 1,795.94 | 1,766.38 | 1,920.91 | 2,107.99 | 2,090.82 | 1,884.04 | **1,922.20** | 1,856.50 |
| Drug 10 MSE | 1,560.88 | 1,599.48 | 1,776.79 | 1,608.28 | 1,608.88 | 1,623.30 | 1,918.77 | 1,795.48 | 1,670.26 | 1,952.52 | **1,711.42** |

**Table 16:** MSE by sub-group exposed to only one of the top-10 RxNorm concepts to evaluated the effects of drug-specific data augmentation of 10% of the total dataset size with a conditional WGAN. Same control on the synthetic time series as with the real data.

| Drug augmented | Baseline real | Drug 1 | Drug 2 | Drug 3 | Drug 4 | Drug 5 | Drug 6 | Drug 7 | Drug 8 | Drug 9 | Drug 10 |
| --- | --- | --- | --- | --- | --- | --- | --- | --- | --- | --- | --- |
| Drug 1 MSE | 1,881.12 | **1,866.19** | 1,791.97 | 2,033.32 | 1,855.53 | 1,790.85 | 1,872.60 | 1,841.75 | 1,853.63 | 1,885.87 | 1,862.61 |
| Drug 2 MSE | 512.87 | 532.91 | **574.57** | 530.07 | 556.31 | 523.31 | 551.25 | 541.77 | 545.32 | 548.28 | 554.04 |
| Drug 3 MSE | 2,328.27 | 2,329.85 | 2,317.37 | **2,366.96** | 2,320.50 | 2,322.19 | 2,333.34 | 2,320.55 | 2,314.10 | 2,328.57 | 2,315.66 |
| Drug 4 MSE | 1,885.59 | 1,897.27 | 1,994.32 | 1,915.85 | **1,842.45** | 1,973.05 | 1,980.87 | 1,894.75 | 1,891.23 | 1,990.13 | 1,927.55 |
| Drug 5 MSE | 1,205.86 | 1,173.76 | 1,170.34 | 1,161.00 | 1,171.23 | **1,219.20** | 1,159.31 | 1,162.27 | 1,178.10 | 1,189.38 | 1,155.37 |
| Drug 6 MSE | 1,036.85 | 972.19 | 927.29 | 919.17 | 987.57 | 955.13 | **1,097.77** | 866.83 | 955.31 | 939.65 | 995.71 |
| Drug 7 MSE | 1,405.50 | 1,457.94 | 1,414.90 | 1,405.32 | 1,420.01 | 1,430.91 | 1,469.95 | **1,520.72** | 1,441.98 | 1,475.09 | 1,474.01 |
| Drug 8 MSE | 1,067.05 | 1,048.53 | 1,111.70 | 1,132.99 | 1,034.31 | 1,111.77 | 1,087.08 | 996.40 | **1,089.39** | 1,033.34 | 1,116.34 |
| Drug 9 MSE | 1,691.60 | 1,698.87 | 1,630.49 | 1,841.07 | 1,729.04 | 1,754.93 | 1,741.81 | 1,743.03 | 1,741.48 | **1,691.10** | 1,658.90 |
| Drug 10 MSE | 1,560.88 | 1,525.10 | 1,542.12 | 1,528.84 | 1,523.18 | 1,526.03 | 1,531.12 | 1,544.17 | 1,531.96 | 1,522.23 | **1,524.60** |

**Table 17:**
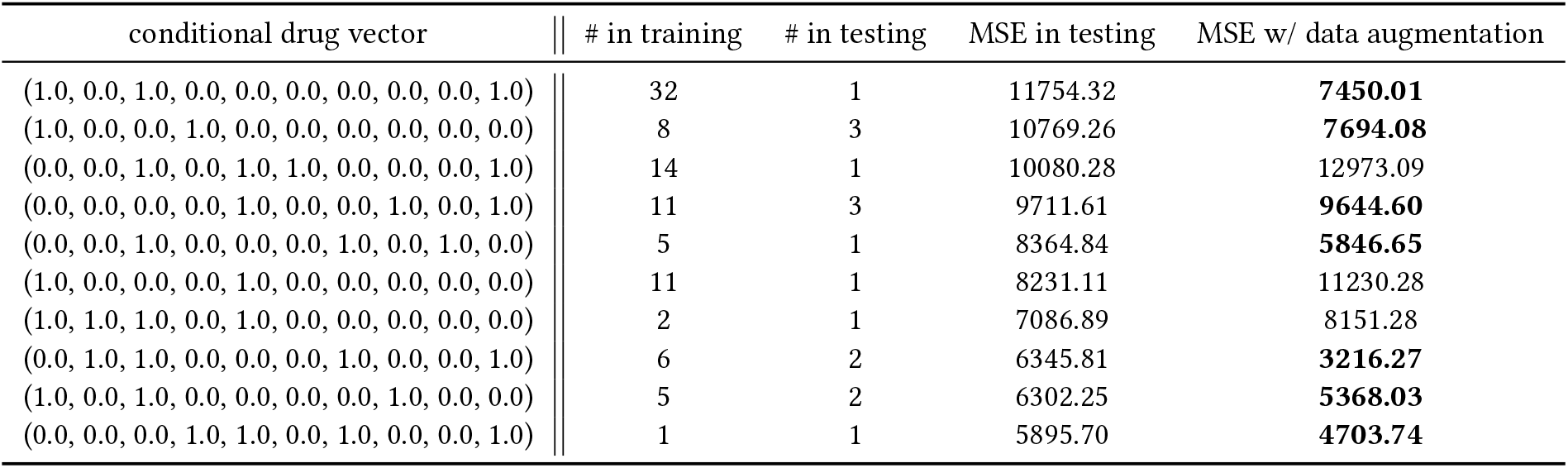
MSE before and after targeted augmentation of the 10 drug combinations with the highest MSE.

## 4 Discussion

In this study, we demonstrated that we can build generative adversarial networks able to generate synthetic laboratory test time series that look like real data. we compared two different models: a Wasserstein GAN with gradient penalty (WGAN-GP) to model regular glucose lab time series, and a conditional WGAN-GP that uses the drug exposure representation introduced in the previous aim to learn better models, but also to control the drug exposure of the synthetic generated time series.

We used two different quantitative methods to evaluate these models: an intrinsic metric called the Fréchet Inception Distance (FID) that consists of measuring the different of the mean and covariance between the synthetic data and the real data; and an extrinsic evaluation that uses the forecasting models developed in the previous aim to evaluate how well models trained on synthetic data only perform on never seen before testing sets.

We first studied the properties of the FID by comparing how the amount of generated data impact the metric when it’s computed with a fixed size training set the GAN was trained on. This experiment showed that the generated data are very stable in spite of the stochasticity of the implicit generative process. We then computed the FID and it’s two components (i.e., the mean and the covariance differences) across epochs to see how they evolve. It seems that most of the time the covariance difference is the main driver of the FID, and that the FID could be a good tool to visualize overfitting of a WGAN model where the FID starts going back up.

We also added visual inspection of the data using the density heatmaps we have been generating throughout the experiments, to help understand how the density learned by the WGAN evolves throughout epochs. It is important to note that all the GAN models developed in this aim had, in spite of their similar mean and standard deviation densities, longer tails. These models generated more extreme values, including some negative measurements (i.e., unrealistic). It exposes one of the main limitation of the Frechet distance that approximates Gaussians and is less sensitive to these longer tails than kernel-based metrics such as maximum mean discrepancy (MMD). Similarly to the forecasting model, these GANs could also benefit from clinically relevant intrinsic evaluation metrics, for instance comparing how real and synthetic data stand in terms of normal ranges, or involving physicians in a qualitative expert evaluation.

The TSTR evaluation demonstrated that the covariance of the synthetic data has an enormous weight on the generalizability of models trained on them. It is also important to note that the performances obtained with synthetic data from conditional models are on-par with the performances of models trained on real data, with an increased MSE that stays within performances obtained with classic machine learning regression models.

Finally, while ATC-3 was a good drug representation for regression, RxNorm appears to be the best conditional information to generate more realistic and generalizable synthetic glucose lab time series. More importantly conditional GANs outperformed the non-conditional model in spite of having to learn a more complex representation of the laboratory test time series.

Only one type of deep generative models, namely generative adversarial networks (GANs) was used. They are not the only types of deep generative models that could have been used. While GANs are implicit models, Variational Autoencoders (VAEs), a type of prescribed model, represent an alternative. Further investigation could be done using VAE, and hybrid models called VAE-GAN [40]. Moreover, supervised models using LSTM have been the standard for sequence generation[24], where text can be generated token by token by sampling from a distribution conditioned on the previous token and a hidden representation of all the tokens already generated. However there are known issues where the models have to sample from conditional distributions never seen at training and their solutions lead to models with no cost function specifically designed to encourage synthetic data fidelity.[35, 20] The GAN models of this aim present the advantage of being self-supervised with a cost function, the Wasserstein distance, designed to explicitly compute an estimate of the distance between real and synthetic samples.

Other auxiliary data could be used as conditional information, such as demographics (i.e., sex, age, race/ethnicity) as they are known to have a direct impact of the laboratory tests distribution and dynamics. Such as conditional WGAN generating laboratory test time series based on demographics, drug exposures, and other clinical covariates could be a module in a larger analysis where these covariates are handpicked by the user for specific generation, or generated themselves stochastically using discrete GANs. With an increasing number of clinical variables, and therefore an increased sparsity, methods to compress patient representations could be used to improve the learning with large auxiliary datasets.

After having evaluated our generative models, we then showed the potential applications for conditional WGAN that can generate irregular glucose time series based on a conditional vector of drug exposures. The two avenues investigated were the inference ability of these generative models, directly tied to arithmetic properties in the latent space of the generator network, and data augmentation in supervised tasks.

In the first experiment, one of the main limitation is the conditional WGAN used, along with the drugs of exposure. This model was selected based on its overall performances at generating synthetic samples close to the real data, and using the Frechet Inception distance. The FID is a good metric to get a sense of the distance between gaussians approximated on two datasets, but not fine grained enough for more thorough comparisons. This is where the maximum mean discrepancy (MMD) metric, a kernel based distance, could be useful. Moreover, there are only 10 drugs in the auxiliary information vector while this population was exposed to hundreds of them. Adding drugs to the auxiliary vector, along with other clinical covariates as discussed in the previous chapter would yield to a better conditional WGAN and better inference. However, most of the inferred time series sub-groups were very close to the data generated by the WGAN that was exposed to them at training.

In the data augmentation task, there was an obvious limitation with the sample size of the time series groups in the testing set. A lot of them had only one sample, resulting in a very noisy and sensitive MSE. Another difficulty comes from the fact that the task is a regression task. Every data augmentation study that has used GANs to improve supervised learning results was applied to classification task. We do think that classification tasks are more robust to data augmentation than regression, due to the more discrete process of selecting a decision threshold, versus a continuous non-linear relationship between inputs and output in regressions. It would also be beneficial to compare the GAN-powered data augmentation with more classic data augmentation methods as a baseline of comparison.

Another limitation of this study was that only one laboratory test, blood glucose, was investigated. However it opens the way for subsequent studies to systematize the methods presented here and refine them.

## 5 Conclusion

The model evaluation tier of this paper is the proof of concept that we can generate laboratory test time series from visits with generative adversarial networks. These synthetic time series are close to the real ones, and yield on-par results when used to train forecasting models then tested on real data never seen by the GAN. The conditional WGAN demonstrated two properties: conditional GANs generate higher quality samples than non-conditional GANs, and they enable the targeted generation of synthetic laboratory test time series exposed to drugs defined at the input of the generator.

With regards to the applications, we demonstrated useful properties of conditional WGAN to simulate drug exposures on laboratory test time series. We showed that these generative models can infer samples behavior based on latent space arithmetic when there is enough useful conditional information available. We also showed that these simulated data can be used in specific tasks where sub-groups of samples are under-powered, causing high errors in these sub-groups. While these studies are only proof of concepts, they show promising applications that could directly impact how researchers work with medical data, and provide the bases for next generation clinical decision making tools that would be able to simulate population specific laboratory test time series.

## Data Availability

The study was conducted with electronic health records (EHR) data from Columbia University Irving Medical Center (CUIMC) and is not publicly available for HIPAA compliance.

